# Pleiotropic and Distributed Neuropsychiatric Effects of Neurodevelopmental Copy Number Variants in the All of Us Biobank

**DOI:** 10.64898/2026.09.13.26362959

**Authors:** Anne Marie Wells, Feiyang Zhao, Sudha Seshadri, Jose Cavazos, Agustin Ruiz

## Abstract

**Background:** Neurodevelopmental copy number variants (ND-CNVs) are associated with diverse neuropsychiatric outcomes, but most evidence derives from clinically ascertained cohorts enriched for severe disease. Whether these associations generalize to large, heterogeneous population biobanks, and whether they reflect locus-specific or distributed genetic effects, remains unclear.

**Methods:** We developed a scalable analytic pipeline within the All of Us Research Program to identify ND-CNV carriers from structural variant callsets and evaluate associations across neuropsychiatric phenotypes derived from electronic health records (EHRs). Logistic regression models estimated associations between carrier status and individual phenotypes, adjusting for age, healthcare utilization, observation time, and genetic ancestry principal components. Sensitivity analyses included exclusion of mosaic chromosomal alteration (mCA)-suspected events and leave-one-locus-out (LOLO) models. Phenotypes were further grouped into Research Domain Criteria (RDoC) domains to assess domain-level structure and cross-domain burden.

**Results:** ND-CNV carrier status was associated with modest but consistent enrichment across neuropsychiatric phenotypes spanning affective, psychotic, neurodevelopmental, and trauma-related domains. Although no individual phenotype survived false discovery rate correction, global permutation analyses demonstrated directional enrichment exceeding expectations under randomized exposure assignment. Effects were distributed rather than driven by a single phenotype or locus, with deletion-overlapping carriers showing stronger associations than duplication-overlapping carriers. Associations remained stable after mCA exclusion and LOLO analyses. Domain-level analyses revealed correlated enrichment across RDoC systems, while cross-domain burden measures demonstrated strong interdependence but limited discriminative separation.

**Conclusions:** ND-CNV carriers exhibit distributed, pleiotropic neuropsychiatric risk in a large biobank. These findings provide empirical support for a distributed model of ND-CNV-associated neuropsychiatric liability in which numerous modest, directionally concordant phenotype associations collectively contribute to pleiotropic risk across neuropsychiatric domains.

## INTRODUCTION

Neurodevelopmental copy number variants (ND-CNVs) represent a class of rare structural variants that confer substantial risk for a wide range of neuropsychiatric outcomes^1–11^. Over the past two decades, studies of recurrent deletions and duplications have demonstrated associations with disorders spanning autism spectrum disorder, schizophrenia, bipolar disorder, and intellectual disability, as well as broader cognitive and functional impairments^5–7,12–17^. These findings have established ND-CNVs as major contributors to psychiatric genetics and as key entry points for understanding the biological basis of neurodevelopmental disorders.

A defining feature of ND-CNVs is their pleiotropy^18–21^. Individual loci often influence multiple neuropsychiatric phenotypes^22^, and carriers frequently exhibit overlapping constellations of symptoms rather than discrete diagnostic presentations^2,23–26^. This pattern is consistent with broader evidence that psychiatric and neurodevelopmental traits share substantial genetic architecture and are not organized as independent disease entities^21,27^. At the biological level, convergent effects of diverse ND-CNVs on synaptic function, neurodevelopmental processes, and large-scale brain networks have been proposed as mechanisms underlying this shared liability^1,4,10,15,18,21,28,29^. Together, these observations support a model in which ND-CNVs act not as locus-specific drivers of single disorders, but as perturbations of interconnected neurobiological systems that manifest across multiple phenotypic domains^30–33^.

Despite these advances, most evidence for ND-CNV-associated neuropsychiatric risk derives from clinically ascertained cohorts enriched for severe or early-onset disease ^6,12–14,34^. While such studies have been essential for locus discovery and characterization, they may overrepresent extreme phenotypes^24,35^ and limit inference about the broader distribution of ND-CNV-associated effects in the general population. Large-scale population biobanks like the U.S.-based *All of Us*^36^ Research Program (AoU) provide an opportunity to address this limitation by enabling the study of ND-CNV carriers across a wide spectrum of clinical presentations and ascertainment contexts. However, identifying biologically meaningful signal in these settings is challenging, given heterogeneous phenotyping, modest effect sizes, and substantial correlation among psychiatric outcomes. Importantly, *All of Us* was not designed as a neuropsychiatric ascertainment cohort, providing an opportunity to evaluate ND-CNV-associated signal outside of disorder-enriched sampling frameworks.

An additional challenge lies in how neuropsychiatric phenotypes are represented analytically. Traditional case–control frameworks centered on single diagnoses may be poorly suited to capturing the distributed and overlapping nature of ND-CNV-associated risk^23,24,37–42^. Dimensional and domain-based approaches, such as the Research Domain Criteria (RDoC) framework^43,44^, may provide a complementary framework for representing distributed and cross-diagnostic neuropsychiatric liability. Coupled with analytic strategies that account for correlated outcomes and cross-domain burden, these approaches may better reflect the structure of genetic and transdiagnostic liability in population-scale data^27,44–47^.

Here, we develop and apply a scalable analytic framework within the AoU Biobank to evaluate ND-CNV-associated neuropsychiatric risk in a large, heterogeneous biobank cohort. Using curated genomic intervals, overlap-based carrier definitions, and electronic health record (EHR)-derived phenotypes, we assess associations at the level of individual neuropsychiatric phenotypes, RDoC-informed domains, and cross-domain burden. We further evaluate robustness through matched control intervals, alternative overlap definitions, and sensitivity analyses addressing ascertainment and locus composition. This framework is designed to evaluate whether heterogeneous ND-CNV loci produce distributed and correlated neuropsychiatric effects consistent with prior models of shared neurobiological and systems-level vulnerability (**Fig. 1**).

**Figure 1.**
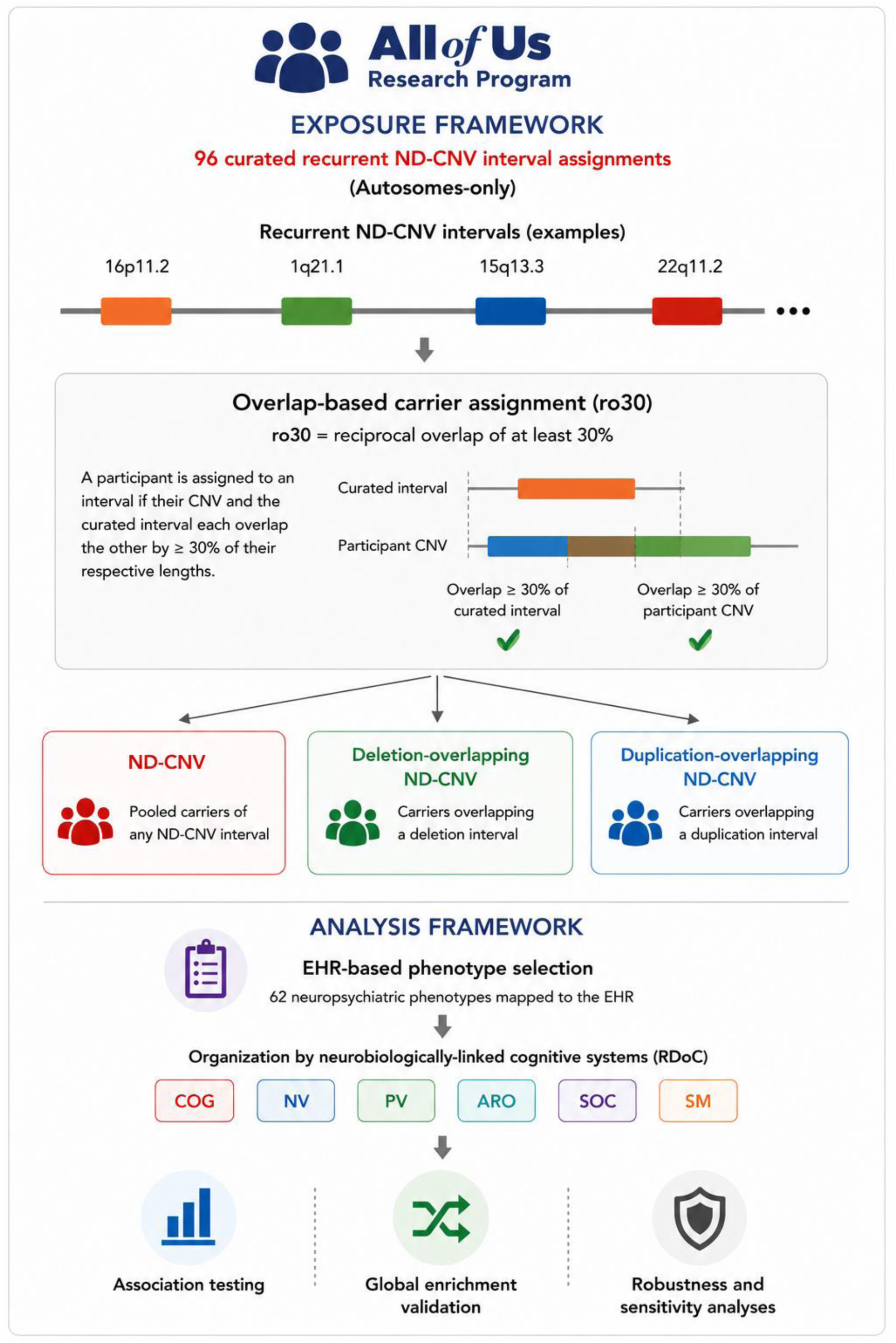
Exposure assignment and analytical framework for evaluation of recurrent neurodevelopmental copy number variants (ND-CNVs) in All of Us. Curated recurrent autosomal ND-CNV intervals (96 interval assignments representing 51 unique loci) were identified and used to define carrier status within the All of Us Research Program. Under the reciprocal-overlap (ro30) assignment strategy, participants were assigned to a curated interval when both the participant CNV and the curated interval overlapped by at least 30% of their respective lengths. The ro30 framework was selected as the primary exposure definition following comparison with multiple alternative overlap thresholds and assignment strategies in sensitivity analyses. This framework generated three analytic exposure models: pooled ND-CNV carriers (ND-CNV), deletion-overlapping ND-CNV carriers (ND-DEL), and duplication-overlapping ND-CNV carriers (ND-DUP). Neuropsychiatric phenotypes derived from electronic health record data (n = 62) were organized into six Research Domain Criteria (RDoC)-informed domains: Cognitive Systems (COG), Negative Valence Systems (NV), Positive Valence Systems (PV), Arousal/Regulatory Systems (ARO), Social Processes (SOC), and Sensorimotor Systems (SM). Associations between ND-CNV exposure status and neuropsychiatric phenotypes were evaluated using phenotype-level regression analyses, followed by global enrichment validation using permutation-based testing and a series of robustness and sensitivity analyses.

## RESULTS

### Analytic cohort and ND-CNV carrier definition

Neurodevelopmental copy number variant (ND-CNV) carrier status in the AoU database was defined using overlap-based matching to curated ND-CNV loci previously identified to carry neuropsychiatric and cognitive deficit risks in large studies^5,12,13,17^. A curated ND-CNV interval set and matched control intervals were constructed to closely match key genomic characteristics, including interval length, GC content, and segmental duplication burden (**Fig. S1**). Matched control intervals showed near-identical distributions across these features relative to ND-CNV loci, supporting their use as a specificity control for distinguishing biologically meaningful signal from background properties of structurally complex genomic regions. Carrier status was analyzed in both pooled and direction-specific frameworks. During pipeline development, alternative overlap definitions—including reciprocal overlap 30% (ro30), ro50, ro70, midpoint and fractional locus-based thresholds—were systematically evaluated (**Fig. S2**). Increasing overlap stringency progressively reduced retained ND-CNV carrier counts across recurrent loci, consistent with exclusion of minimally overlapping or structurally heterogeneous events (**Fig. S2A**). External benchmarking against the gnomAD-SV v2.1 population structural variant resource demonstrated broad contextual concordance between AoU recurrent ND-CNV prevalence estimates and overlapping population SV frequencies, while also illustrating the expected reduction in matched overlapping events under stricter reciprocal-overlap thresholds (ro50) relative to the primary ro30 framework (**Fig. S2B**).

Thus, the primary analytic cohort comprised 71,992 participants. In domain-level models, this corresponded to 2,115 ND-CNV carriers overall, including 1,005 deletion-overlapping ND-CNV carriers and 1,786 duplication-overlapping ND-CNV carriers. Because participants may carry both deletion-overlapping and duplication-overlapping ND-CNV assignments, these exposure counts are not mutually exclusive and should be interpreted as model-specific exposure groups rather than distinct participant cohorts. Cohort composition, exposure structure, and ancestry distributions demonstrated broad representation of ND-CNV carriers across genetic ancestry groups and supporting inclusion of ancestry principal components together with healthcare-utilization and observation-time covariates in downstream models. (**Fig. S3**).

To evaluate whether observed associations reflected locus-specific biological effects rather than general properties of structural variation, ND-CNV exposure was compared to matched control interval exposure. These control intervals were selected from genomic regions outside the curated ND-CNV locus set and were matched to ND-CNV loci on chromosome, interval length, GC content, and segmental duplication burden, thereby providing a genomic background control for structural variation independent of established neurodevelopmental risk loci. While ND-CNV carrier status showed distributed nominal enrichment across neuropsychiatric phenotypes, matched control intervals yielded largely null or inconsistent estimates across the same models (**Fig. S3C**). This divergence supported the specificity of the ND-CNV signal and argued against confounding by genomic architecture or variant burden alone.

### Distributed neuropsychiatric enrichment among ND-CNV carriers

Neuropsychiatric phenotypes were constructed using a dictionary-driven framework that integrated standardized OMOP concept expansion defining instance of neuropsychiatric symptoms and/or disorders within participants’ electronic health records data. Then, in effort to organize neuropsychiatric symptoms by underlying functional systems rather than traditional diagnostic categories, OMOP concepts were mapped to Research Domain Criteria (RDoC)–informed domains (**Fig. S4**). In total, 62 **phenotypes** were defined and grouped into six primary RDoC domains—social processes, sensorimotor systems, arousal/regulatory systems, negative valence systems, cognitive systems, and positive valence systems, permitting representation of overlapping and cross-domain clinical features.

Across individual phenotype-level analyses, pooled ND-CNV carriers demonstrated nominal enrichment across multiple neuropsychiatric domains (**Fig. 2A**). The strongest association was observed for anhedonia (OR = 5.55 [95% CI: 2.08–14.84], P = 4.07 × 10⁻⁴), with additional nominal associations observed for posttraumatic stress disorder (OR = 1.37 [95% CI: 1.13–1.66], P = 0.0011), paranoid schizophrenia (OR = 2.04 [95% CI: 1.25–3.34], P = 0.0047), schizophrenia (OR = 1.54 [95% CI: 1.13–2.11], P = 0.0057), mania (OR = 2.15 [95% CI: 1.17–3.96], P = 0.014), and several related psychotic and cognitive phenotypes including delusions, delirium, and confusional state. Additional nominal associations were observed for tremor and repetitious behavior. Although no individual phenotype survived false discovery rate (FDR) correction, the overall pattern was notable for its breadth across affective, psychotic, cognitive, and sensorimotor domains.

**Figure 2.**
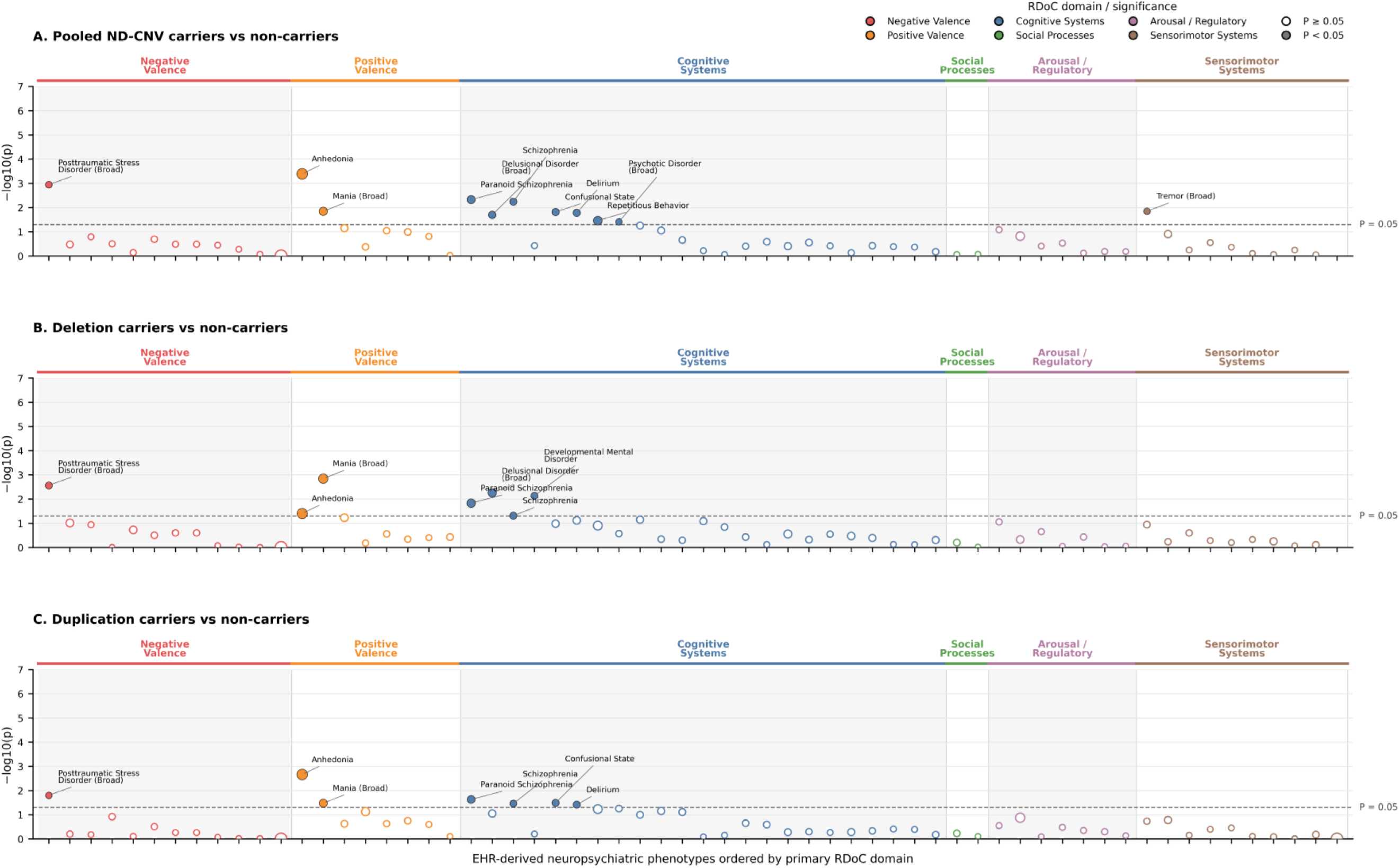
Phenome-wide architecture of neurodevelopmental copy number variant (ND-CNV) associations across neuropsychiatric phenotypes in All of Us. PheWAS-style Manhattan plots showing associations between ND-CNV exposure status and neuropsychiatric phenotypes organized by primary Research Domain Criteria (RDoC) domain. (A) Pooled ND-CNV carriers versus non-carriers, (B) deletion-overlapping ND-CNV carriers versus non-carriers, and (C) duplication-overlapping ND-CNV carriers versus non-carriers. Each point represents a phenotype-specific logistic regression model adjusted for age at last observation, healthcare utilization (log-transformed number of unique clinical visits), observation time, and ancestry principal components (PC1–PC5). The y-axis displays association significance as −log10(P), while point size is proportional to the magnitude of the estimated effect size (|log₂ odds ratio|). Colors indicate primary RDoC domain membership. Filled points denote nominally significant associations (uncorrected P < 0.05), whereas open points denote non-significant associations (P ≥ 0.05). Phenotypes are ordered by primary RDoC domain and secondarily by effect size within domain. Annotated phenotypes represent nominally significant associations. The horizontal dashed line indicates the nominal significance threshold (P = 0.05). Overall, pooled, deletion-overlapping, and duplication-overlapping ND-CNV exposures demonstrate distributed patterns of neuropsychiatric association, with stronger and more numerous nominal associations observed among deletion-overlapping carriers.

Deletion-overlapping ND-CNV carriers demonstrated the strongest and most consistent pattern of neuropsychiatric enrichment (**Fig. 2B**). Nominal associations were observed for mania (OR = 3.20 [95% CI: 1.56–6.57], P = 0.0014), posttraumatic stress disorder (OR = 1.50 [95% CI: 1.15–1.94], P = 0.0027), delusional disorder (OR = 2.20 [95% CI: 1.26–3.85], P = 0.0056), developmental mental disorder (OR = 1.47 [95% CI: 1.11–1.95], P = 0.0073), paranoid schizophrenia (OR = 2.31 [95% CI: 1.18–4.53], P = 0.0148), anhedonia (OR = 4.50 [95% CI: 1.07–18.86], P = 0.0396), and schizophrenia (OR = 1.56 [95% CI: 1.00–2.42], P = 0.0490). These findings spanned positive valence, negative valence, cognitive, and developmental domains, consistent with a broadly distributed pattern of neuropsychiatric liability.

Duplication-overlapping ND-CNV carriers demonstrated a more heterogeneous but directionally similar pattern of associations (**Fig. 2C**). The largest effect estimate was again observed for anhedonia (OR = 5.14 [95% CI: 1.80–14.65], P = 0.0022), with additional nominal associations observed for posttraumatic stress disorder (OR = 1.30 [95% CI: 1.05–1.61], P = 0.0157), paranoid schizophrenia (OR = 1.92 [95% CI: 1.09–3.36], P = 0.0234), confusional state (OR = 1.57 [95% CI: 1.04–2.38], P = 0.0322), mania (OR = 2.08 [95% CI: 1.06–4.08], P = 0.0332), schizophrenia (OR = 1.46 [95% CI: 1.03–2.06], P = 0.0346), and delirium (OR = 1.56 [95% CI: 1.03–2.37], P = 0.0384). While duplication-overlapping carriers generally exhibited fewer nominal associations and somewhat smaller effect estimates than deletion-overlapping carriers, the overall pattern remained broadly consistent across affective, psychotic, and cognitive phenotypes.

At the domain level, aggregation of phenotype-level effects revealed modest and consistently positive associations across RDoC domains (**Fig. S5**), with similar directional patterns observed across pooled, deletion-overlapping, and duplication-overlapping exposure models. Direct comparison of phenotype-level effect estimates further supported this pattern, with deletion-overlapping models showing larger effects than duplication-overlapping models across a greater proportion of evaluated phenotypes (**Fig. S6**). Together, these findings support a distributed and pleiotropic pattern of ND-CNV-associated neuropsychiatric liability spanning multiple functional domains.

### Positive-control benchmarking confirms expected ND-CNV-associated signal

To evaluate whether the analytic framework recapitulates known ND-CNV-associated phenotypes, we examined a curated set of neurodevelopmental and psychiatric positive-control outcomes (**Fig. 3**). Among pooled ND-CNV carriers, nominal enrichment was observed across multiple expected phenotypes (**Fig. 3A**), with the strongest effects seen in schizophrenia (OR = 1.54 [95% CI: 1.13-2.10], *P* = 0.0057), mania (OR = 2.16 [95% CI: 1.17-3.98], *P* = 0.0144) and psychosis-related outcomes. Additional directional increases were observed across developmental and seizure-related phenotypes, although effect sizes were generally modest and not all associations reached nominal significance.

**Figure 3.**
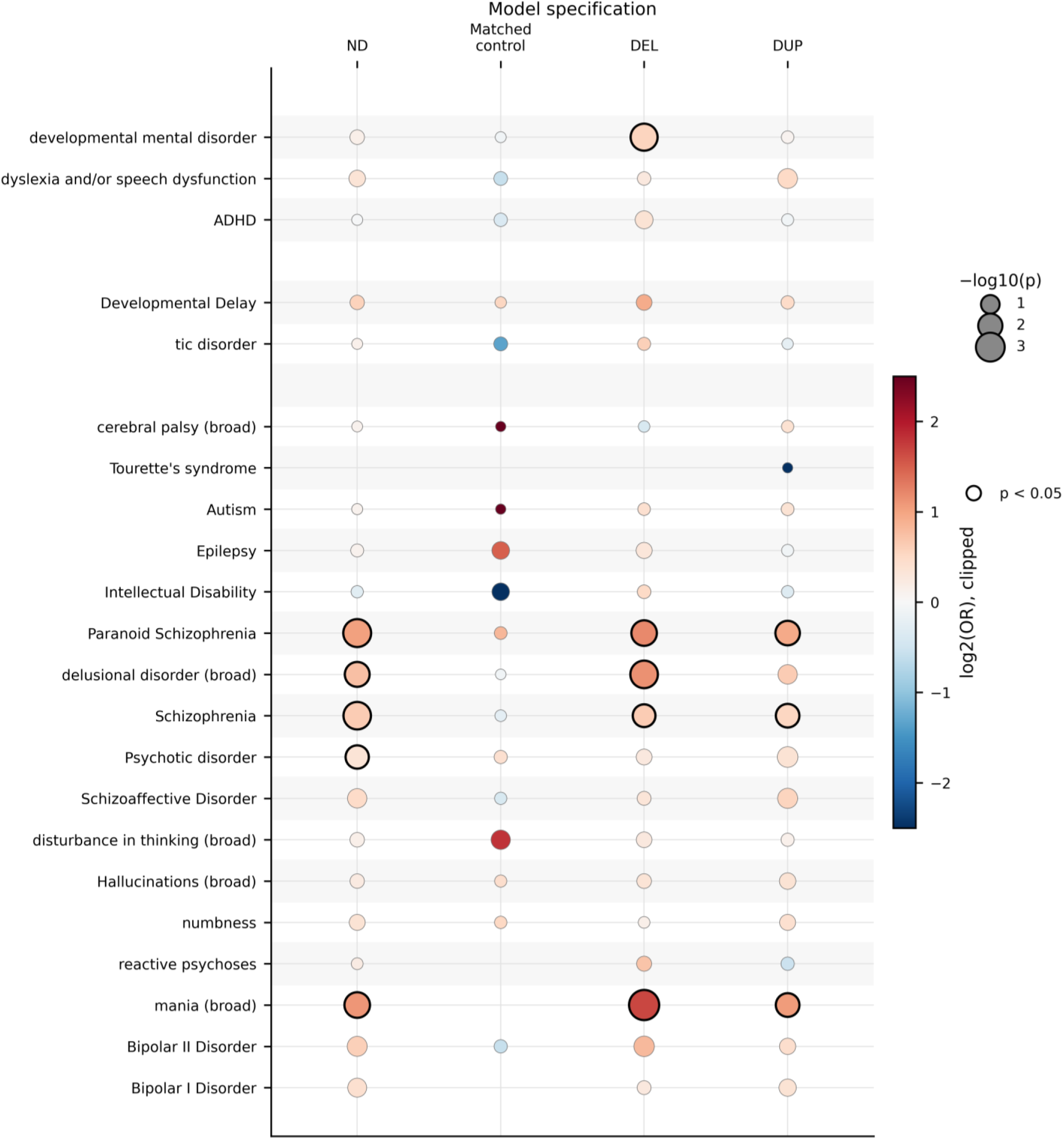
Positive-control benchmarking of expected neurodevelopmental and psychiatric phenotypes among ND-CNV carriers. Dot-matrix visualization of associations between ND-CNV exposure status and a curated set of neurodevelopmental and psychiatric phenotypes previously reported in ND-CNV cohorts. Rows represent benchmark phenotypes and columns represent analytic models: pooled ND-CNV carriers versus non-carriers (ND), matched control interval carriers versus non-carriers (Matched Control), deletion-overlapping ND-CNV carriers versus non-carriers (DEL), and duplication-overlapping ND-CNV carriers versus non-carriers (DUP). Dot color represents effect size (log₂ odds ratio), with warmer colors indicating increased risk and cooler colors indicating reduced risk. Dot size is proportional to association significance (−log10[P]). Black outlines indicate nominally significant associations (uncorrected P < 0.05). All models were adjusted for age at last observation, healthcare utilization (log-transformed number of unique clinical visits), observation time, and ancestry principal components (PC1–PC5). The overall pattern recapitulates established ND-CNV-associated neuropsychiatric liability across developmental, psychotic, and mood-related phenotypes. Stronger enrichment among pooled ND-CNV and deletion-overlapping exposures, coupled with attenuation among matched control intervals, supports the biological specificity of the observed signal and the validity of the analytic framework.

When compared against carriers of matched control intervals, effect estimates were attenuated and less consistent across phenotypes (**Fig. 3B**), with wider confidence intervals and reduced statistical power, though the overall direction of effect remained broadly concordant with the primary analysis. Deletion-overlapping ND-CNV carriers demonstrated the strongest and most consistent enrichment across positive-control phenotypes (**Fig. 3C**). Elevated odds were observed across bipolar/mania phenotypes and multiple psychotic disorders, including paranoid schizophrenia, delusional disorder, and schizophrenia, with several phenotypes reaching nominal significance. Developmental phenotypes showed more modest and variable effect estimates.

In contrast, duplication-overlapping ND-CNV carriers exhibited weaker and more heterogeneous associations (**Fig. 3D**), with most effect estimates centered closer to the null and fewer nominally significant findings across phenotypes. Together, these results demonstrate that the analytic framework recovers expected, directionally consistent enrichment across established ND-CNV-associated phenotypes, particularly for deletion-overlapping carriers, providing internal validation of the approach. To contextualize these findings relative to prior clinically ascertained and biobank-based ND-CNV studies, we compared locus-level directional enrichment patterns observed in All of Us against previously reported neurodevelopmental and neuropsychiatric associations across recurrent ND-CNV loci. Despite differences in ascertainment strategy, cohort composition, and phenotype definition, AoU-derived association patterns demonstrated broad qualitative concordance with previously established literature signals across multiple loci and phenotypic domains (**Fig. S7**).

### Global permutation analyses support structured neuropsychiatric enrichment beyond individual phenotype associations

Although no individual phenotype-level association survived false discovery rate correction, the aggregate pattern of ND-CNV-associated neuropsychiatric effects demonstrated substantially greater directional consistency than expected under random exposure assignment (**Fig. 4**). To evaluate whether the observed enrichment pattern reflected structured signal rather than isolated nominal associations, we performed a phenotype-level permutation analysis in which ND-CNV exposure labels were randomly reassigned across participants while preserving the observed phenotype matrix and analytic framework.

**Figure 4.**
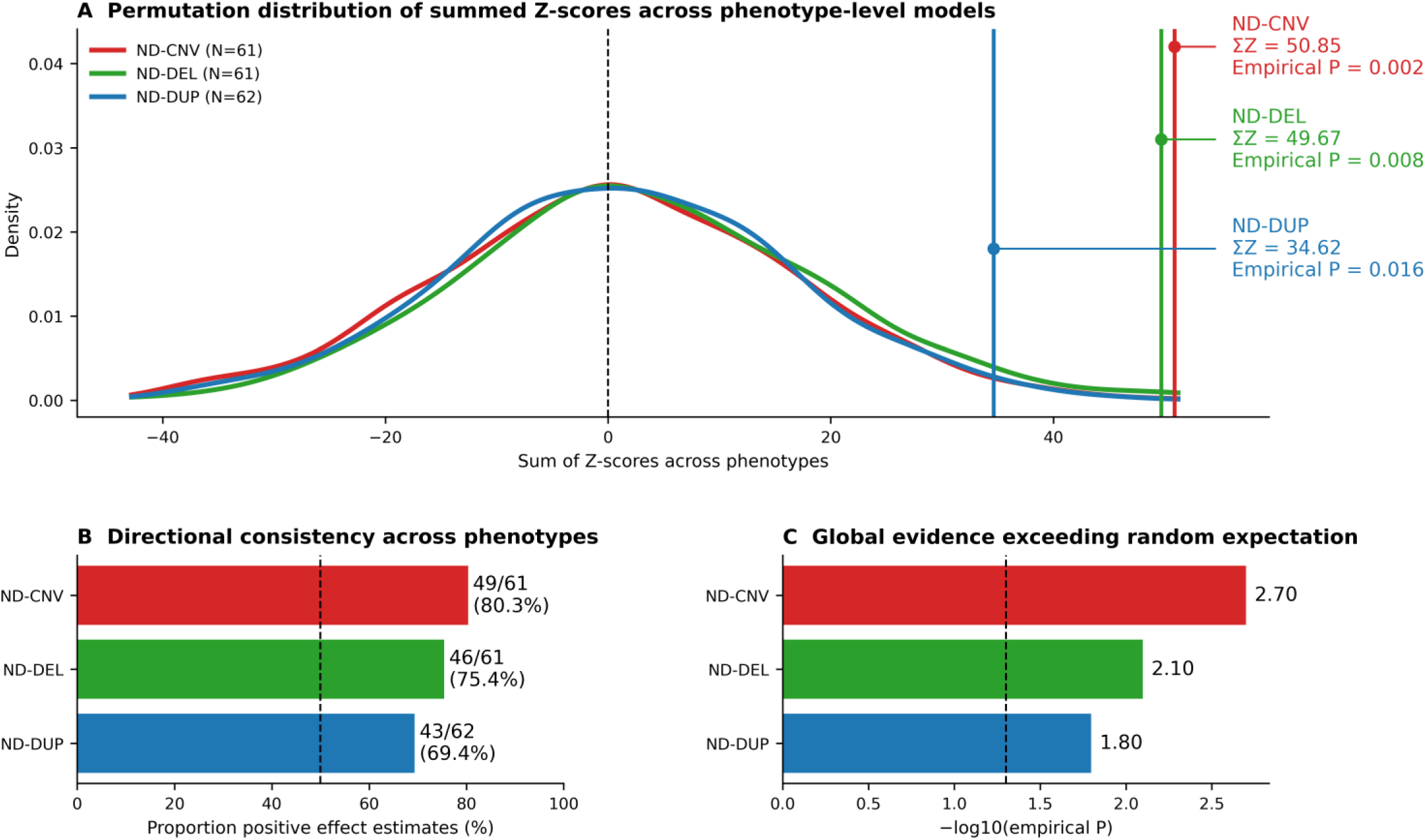
Global permutation analyses support structured neuropsychiatric enrichment among ND-CNV carriers. **(A)** Empirical null distributions of aggregate neuropsychiatric enrichment generated from 1,000 exposure-permutation replicates. For each permutation, ND-CNV exposure labels were randomly reassigned while preserving the observed phenotype structure and analytic framework. Curves represent null distributions of the summed Z-score statistic (ΣZ) across all phenotype-level logistic regression models for pooled ND-CNV carriers (ND-CNV), deletion-overlapping ND-CNV carriers (ND-DEL), and duplication-overlapping ND-CNV carriers (ND-DUP). Vertical lines indicate the observed summed Z-score statistic from the primary analyses. Across all exposure definitions, observed enrichment substantially exceeded expectations under randomized exposure assignment. **(B)** Directional consistency across phenotype-level associations. Bars indicate the proportion of neuropsychiatric phenotypes exhibiting positive effect estimates (β > 0) in the observed analyses. The dashed line denotes the 50% expectation under random assignment. Pooled ND-CNV carriers demonstrated positive effect estimates for 49 of 61 phenotypes (80.3%), deletion-overlapping ND-CNV carriers for 46 of 61 phenotypes (75.4%), and duplication-overlapping ND-CNV carriers for 43 of 62 phenotypes (69.4%). **(C)** Global evidence exceeding random expectation. Bars represent empirical significance of the observed summed Z-score statistic expressed as −log10(empirical *P*). The dashed line indicates the nominal significance threshold (*P* = 0.05). Empirical permutation probabilities were 0.002 for pooled ND-CNV carriers, 0.008 for deletion-overlapping ND-CNV carriers, and 0.016 for duplication-overlapping ND-CNV carriers.

Across 61 neuropsychiatric phenotypes evaluated in the pooled ND-CNV analysis, 49 (80.3%) demonstrated positive effect estimates, substantially exceeding the proportion expected under randomized exposure assignment (**Fig. 4B**). Similarly, aggregate directional enrichment across phenotypes was elevated relative to the empirical null distribution, with the observed summed Z-score statistic (ΣZ = 50.85) exceeding nearly all randomized permutations (empirical P = 0.002; **Fig. 4A,C**). These findings indicate that the overall pattern of ND-CNV-associated neuropsychiatric effects is unlikely to arise through stochastic variation alone.

Direction-specific analyses yielded similar results. Deletion-overlapping ND-CNV carriers demonstrated the strongest aggregate enrichment signal, with 46 of 61 phenotype-level associations (75.4%) exhibiting positive effect estimates and a summed Z-score of 49.67 (empirical P = 0.008). Duplication-overlapping ND-CNV carriers demonstrated a more attenuated but still directionally consistent pattern, with 43 of 62 phenotypes (69.4%) exhibiting positive effect estimates and a summed Z-score of 34.62 (empirical P = 0.016). Across all exposure definitions, observed directional enrichment consistently exceeded expectations derived from randomized exposure assignments.

Importantly, these findings suggest that the observed ND-CNV signal is not driven by a small number of isolated phenotype associations. Rather, numerous neuropsychiatric phenotypes demonstrate modest but directionally coherent effects that collectively produce a distributed enrichment pattern. The permutation analyses therefore provide complementary evidence that the observed neuropsychiatric architecture reflects structured pleiotropic signal rather than random fluctuations arising from multiple testing alone.

Sensitivity analyses using alternative global enrichment metrics yielded highly concordant results (**Fig. S8**). Significant enrichment was observed not only for the primary summed Z-score statistic (ΣZ), but also for a signed enrichment statistic incorporating effect direction and significance as well as for the proportion of phenotypes demonstrating positive effect estimates. The consistency of these findings across multiple enrichment frameworks supports the robustness of the observed ND-CNV-associated neuropsychiatric architecture and argues against dependence on any single summary metric.

### Robustness analyses demonstrate stability of ND-CNV–associated effects

To assess the stability and specificity of ND-CNV–associated neuropsychiatric effects, we performed a series of sensitivity analyses addressing potential confounding by healthcare utilization, somatic variation, locus-specific effects, and assigned ND-CNV locus burden (**Fig. 5**). Adjustment for healthcare utilization and observation time had minimal impact on effect estimates (**Fig. 5A; Fig. S9**). Across all phenotypes assessed, including developmental mental disorder, schizophrenia, and mania, odds ratios were nearly identical between reduced and fully adjusted models, indicating that observed associations are not driven by differential healthcare utilization or diagnostic opportunity. Exclusion of participants harboring variants flagged as potential mosaic chromosomal alterations (mCAs; N = 5 flagged cases) similarly resulted in minimal change in effect estimates (**Fig. 5B**). Associations for key phenotypes—including developmental delay, schizophrenia, and mania—remained stable, with overlapping confidence intervals and no systematic attenuation, supporting robustness to potential blood-derived structural variation artifacts.

**Figure 5.**
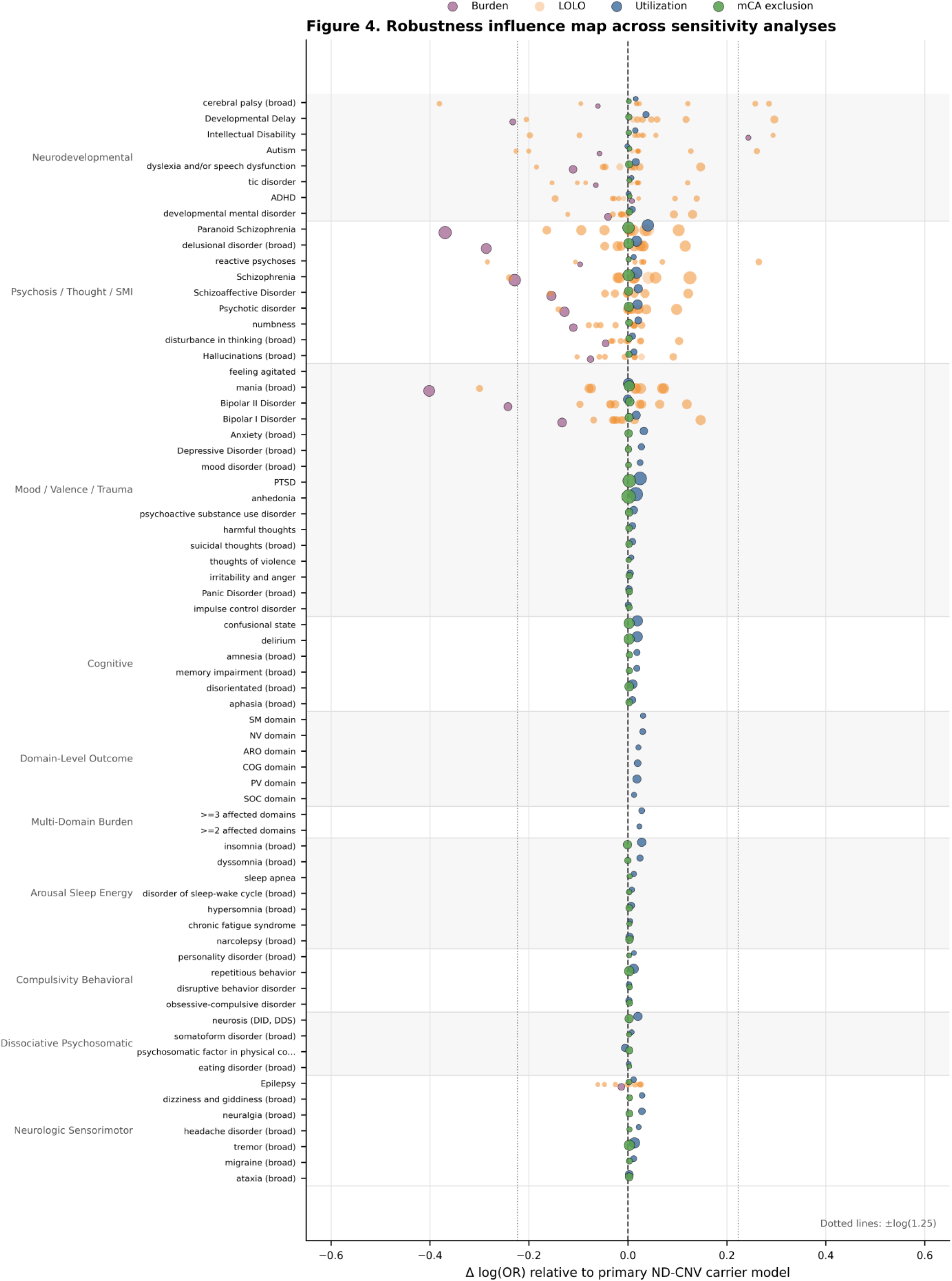
Robustness of ND-CNV associations across sensitivity analyses. Influence-map visualization of the stability of ND-CNV effect estimates across multiple sensitivity analyses. Each point represents a phenotype-specific association evaluated under an alternative analytic framework, including healthcare-utilization adjustment (**UTIL**), mosaic chromosomal alteration exclusion (**mCA**), leave-one-locus-out analyses (**LOLO**), and CNV locus assignment burden modeling (**BURDEN**). The x-axis displays the change in effect size relative to the primary model (Δ log odds ratio), with values near zero indicating minimal deviation from the primary estimate. Point size is proportional to association significance (–log₁₀[P]), and colors denote sensitivity-analysis category. Vertical reference lines indicate no change in effect size relative to the primary model. All sensitivity analyses were performed using the same covariate framework as the primary models unless otherwise specified. The overall clustering of associations around Δ log odds ratio = 0 demonstrates that observed ND-CNV-associated neuropsychiatric signals are robust to alternative model specifications, exclusion of potentially confounding genomic events, individual-locus influence, and differences in overall CNV burden. The absence of systematic directional shifts across sensitivity analyses supports the interpretation that the observed phenotypic architecture reflects distributed ND-CNV-associated liability rather than ascertainment artifacts, individual high-impact loci, or model-specific effects.

Leave-one-locus-out (LOLO) analyses demonstrated that the overall pattern of ND-CNV-associated neuropsychiatric enrichment remained broadly preserved following exclusion of individual recurrent loci, although several loci produced more substantial deviations than others (**Fig. 5C**; extended results **Fig. S10-11**). Exclusion of 2q13 (NPHP1) and 15q13.3 BP4-BP5 produced the largest reductions in global enrichment metrics (Δ mean Z = −0.377 and −0.327, respectively), whereas exclusion of 15q11q13 BP3-BP5 also substantially attenuated enrichment (Δ mean Z = −0.271). Locus-specific association profiles demonstrated that 2q13 (NPHP1) contributed to multiple schizophrenia-spectrum and developmental phenotypes, including schizophrenia, psychotic disorder, paranoid schizophrenia, schizoaffective disorder, and dyslexia/speech dysfunction, while 15q13.3 BP4-BP5 showed strongest associations with mania and developmental mental disorders (**Fig. S11B**). In contrast, exclusion of several less prevalent loci, including TAR, 1q21.1, and distal 22q11.2 intervals, produced comparatively modest or localized changes in enrichment metrics. No single locus eliminated the overall pattern of neuropsychiatric enrichment, and most leave-one-locus-out estimates remained directionally concordant with the full ND-CNV model, suggesting that the observed architecture reflects distributed contributions from multiple recurrent ND-CNV loci rather than dependence on a single dominant interval.

Finally, models incorporating assigned ND-CNV locus count yielded effect estimates that were broadly concordant with binary carrier models (**Fig. 5D**). For several phenotypes, including schizophrenia and mania, burden-based models produced modestly larger effect estimates, consistent with increasing neuropsychiatric liability across participants assigned to greater numbers of curated ND-CNV loci. While these findings should be interpreted cautiously given the overlap-based exposure framework, they suggest that the aggregate enrichment signal is not solely attributable to carrier status alone and may scale with the extent of assigned ND-CNV exposure.

### Structured correlation reveals distributed organization of ND-CNV–associated phenotypes

To characterize higher-order organization of ND-CNV–associated phenotypes, we examined correlation structure across RDoC domains and multi-domain burden measures (**Fig. 6**). At the domain level, pairwise correlations were uniformly positive and modest to moderate in magnitude (**Fig. 6A**; r = 0.11–0.38), indicating that neuropsychiatric phenotypes tend to co-occur across domains while retaining domain-specific structure. The strongest correlations were observed between negative valence and arousal/regulatory systems (r = 0.38) and between sensorimotor and negative valence systems (r = 0.36), while other domain pairs showed weaker but consistently positive associations.

**Figure 6.**
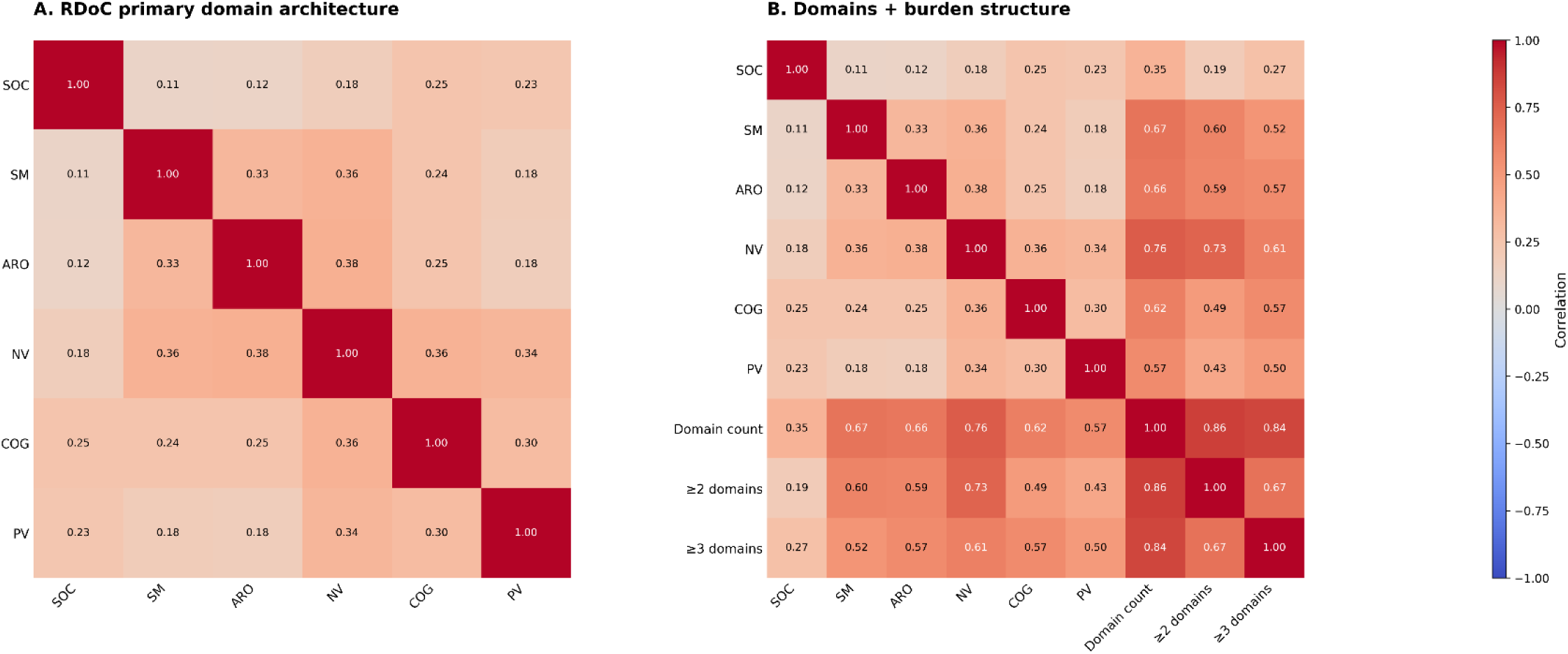
Correlation architecture of RDoC domains and ND-CNV–associated phenotypic burden. Heatmaps display pairwise Pearson correlations among RDoC-informed domain indicators and multi-domain burden measures. Values within cells indicate correlation coefficients, and color intensity reflects correlation magnitude; diagonal elements are fixed at 1.00. Correlations were calculated using Pearson correlation coefficients across domain-level and burden-level phenotype indicators. **(A)** Correlation structure among the six primary RDoC domains: social processes (SOC), sensorimotor systems (SM), arousal/regulatory systems (ARO), negative valence systems (NV), cognitive systems (COG), and positive valence systems (PV). Correlations are uniformly positive and modest to moderate in magnitude (range: r = 0.11–0.38), with the strongest relationships observed between negative valence and arousal/regulatory systems (r = 0.38) and between sensorimotor and negative valence systems (r = 0.36), indicating shared variance across domains while preserving domain-specific structure. **(B)** Correlation structure including both RDoC domains and aggregate burden measures: total domain count, presence of phenotypes spanning ≥2 domains, and presence of phenotypes spanning ≥3 domains. Domain count is strongly correlated with multi-domain burden measures (r = 0.86 for ≥2 domains; r = 0.84 for ≥3 domains). Among individual domains, negative valence shows the strongest association with domain count (r = 0.76), followed by sensorimotor systems (r = 0.67) and arousal/regulatory systems (r = 0.66). The ≥2-domain burden measure is most strongly associated with negative valence (r = 0.73), while ≥3-domain burden remains moderately correlated across all domains (r ≈ 0.50–0.61).

Incorporating aggregate burden measures revealed a complementary pattern (**Fig. 6B**). Total domain count was strongly correlated with multi-domain burden indicators (r = 0.86 for ≥2 domains and r = 0.84 for ≥3 domains), confirming internal consistency of the burden metrics. Among individual domains, negative valence showed the strongest association with domain count (r = 0.76), followed by sensorimotor systems (r = 0.67) and arousal/regulatory systems (r = 0.66). Multi-domain burden measures were also most strongly associated with negative valence (r = 0.73 for ≥2 domains), with moderate correlations observed across all other domains. Importantly, no single domain fully accounted for multi-domain burden, and correlations across domains remained well below unity. This pattern indicates that ND-CNV–associated phenotypic burden reflects distributed involvement across multiple neuropsychiatric systems, rather than concentration within a single domain.

Exploratory pathway enrichment analyses of genes overlapping curated ND-CNV intervals identified broad enrichment patterns across multiple orthogonal annotation frameworks, including GO Biological Process, Reactome, SynGO, Human Phenotype Ontology, and DisGeNET resources (**Fig. 7**). Enriched functional pathways included neurodevelopmental, synaptic, neuronal signaling, and cellular regulatory processes, while clinical annotation enrichment highlighted developmental delay, autism spectrum disorder, ADHD, and related neuropsychiatric phenotypes. Shared-gene network visualization illustrated overlapping gene membership across multiple enriched clinical annotation terms, consistent with distributed and pleiotropic models of ND-CNV-associated neuropsychiatric liability. These analyses are intended primarily as contextual biological annotation rather than definitive mechanistic inference.

**Figure 7.**
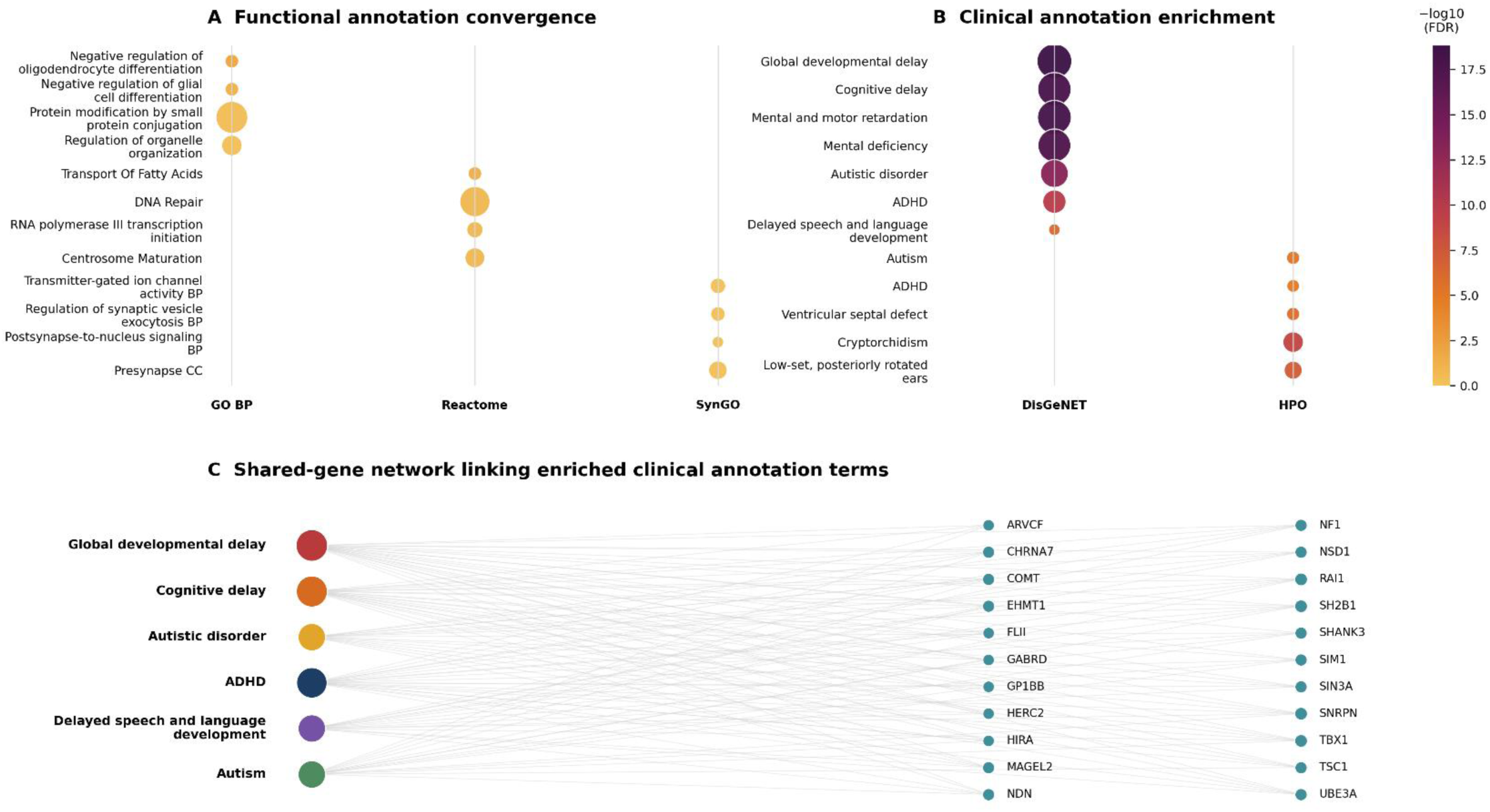
Exploratory pathway convergence among genes overlapping curated ND-CNV intervals. Exploratory pathway enrichment analyses were performed using protein-coding genes overlapping curated neurodevelopmental copy number variant (ND-CNV) intervals identified in the present study. Gene annotations were derived from GENCODE GRCh38 references, and enrichment analyses were conducted using GO Biological Process, Reactome, SynGO, Human Phenotype Ontology (HPO), and DisGeNET resources. Bubble color represents enrichment strength (−log10 FDR-adjusted P value), while bubble size reflects the number of overlapping ND-CNV genes contributing to each enrichment term. **(A)** Functional annotation enrichment across GO Biological Process, Reactome, and SynGO databases identified overrepresentation of pathways related to neurodevelopmental processes, synaptic organization, neuronal signaling, and cellular regulatory mechanisms among genes overlapping curated ND-CNV intervals. **(B)** Clinical annotation enrichment across DisGeNET and HPO resources demonstrated enrichment of neurodevelopmental and neuropsychiatric phenotypes, including developmental delay, autism spectrum disorder, ADHD, cognitive impairment, and related clinical features. **(C)** Shared-gene network linking enriched clinical annotation terms to overlapping ND-CNV genes. Edges represent shared gene membership relationships derived from the enrichment framework and are intended as a visual summary of convergent biological organization rather than a mechanistic interaction model.

## DISCUSSION

In this study, we developed and applied a scalable analytic framework within the All of Us Research Program to evaluate the neuropsychiatric impact of neurodevelopmental copy number variants (ND-CNVs) in a large, heterogeneous EHR-linked population cohort. Using overlap-based carrier definitions, phenotype-level association testing, and RDoC-informed domain mapping, we observed a consistent pattern of modest, distributed enrichment across neuropsychiatric phenotypes. Although no individual association survived FDR correction, the overall structure of results—spanning affective, psychotic, neurodevelopmental, and trauma-related outcomes—was coherent and reproducible across multiple analytic conditions. Importantly, global permutation analyses demonstrated that the aggregate pattern of ND-CNV-associated neuropsychiatric effects substantially exceeded expectations under randomized exposure assignment, indicating that the observed signal reflects structured directional enrichment rather than isolated nominal associations. The central finding is therefore not a single high-confidence association, but evidence of a structured pattern of pleiotropic signal that aligns with the known biology of ND-CNVs^11,21,27,48^. This distinction is important: in a population-scale biobank with heterogeneous ascertainment^36^, the expected manifestation of ND-CNV risk is not a single dominant phenotype, but rather a distributed liability across related neuropsychiatric domains^27,43,44,49–51^.

### Evidence for distributed ND-CNV-associated neuropsychiatric liability

Our results are consistent with a large body of work demonstrating that ND-CNVs confer broad and variable neuropsychiatric risk. Rare structural variants have long been recognized as major contributors to neurodevelopmental and psychiatric disorders, often spanning multiple diagnostic categories rather than mapping cleanly onto a single condition^1,4,15,18,52–54^. Studies in clinically ascertained cohorts and population samples have shown that pathogenic ND-CNVs are associated with schizophrenia, autism, bipolar disorder, cognitive impairment, and general functional outcomes^6,7,12,13,24,26,28,55,56^.

The phenotype-level results observed here recapitulate this pleiotropic architecture. Rather than a single disorder dominating the signal, we observe nominal enrichment across a range of clinically related phenotypes, including psychotic, affective, developmental, and trauma-associated outcomes. This pattern aligns with prior findings that ND-CNV carriers often exhibit overlapping and variably expressed symptom profiles, reflecting shared underlying liability rather than discrete disease categories. Importantly, our results extend these observations into a biobank-scale population, demonstrating that pleiotropic ND-CNV signal can be recovered even in the absence of clinical ascertainment. This suggests that the distributed architecture of CNV-associated risk is not solely an artifact of case-enriched cohorts but reflects a more generalizable feature of their biological effects^6,12,13,18–21^.

### Gene dosage and variable expressivity across recurrent ND-CNVs

Importantly, locus-specific enrichment patterns observed within All of Us demonstrated broad qualitative concordance with previously established ND-CNV literature despite substantial differences in cohort ascertainment, diagnostic structure, and phenotyping methodology. Canonical loci associated with cognitive impairment, schizophrenia-spectrum disorders, neurodevelopmental syndromes, and broader psychiatric burden in clinically enriched cohorts frequently demonstrated directionally consistent enrichment patterns within the AoU EHR environment. Directional analyses provide additional insight into the structure of ND-CNV effects. Across models, deletion-overlapping ND-CNV carriers exhibited somewhat stronger and more diffuse enrichment across phenotypes compared to duplication-overlapping ND-CNV carriers, although both groups showed overlapping patterns of liability. This aligns with prior evidence that gene dosage effects can differ between deletions and duplications, with deletions often producing larger phenotypic shifts^13,15,28,35,57^.

However, the overall similarity between deletion and duplication profiles suggests that genes within these loci contribute to shared dimensions of neuropsychiatric liability. Rather than producing entirely distinct phenotypic spectra, our results suggest that ND-CNV direction appears to modulate the magnitude and distribution of effects within a common underlying liability structure. This aligns with emerging models of variable expressivity, in which genetic modifiers, environmental factors, and ascertainment interact to shape ND-CNV-related liabilities^21^. Thus, comparisons should be interpreted cautiously. The literature-concordance analysis was qualitative and domain-based rather than a formal meta-analytic replication framework, and several locus-domain combinations were subject to privacy-related suppression thresholds within the All of Us environment. Moreover, psychiatric diagnoses derived from EHR systems represent observed clinical coding patterns rather than definitive absence-or-presence determinations, particularly for underdiagnosed or variably documented neurodevelopmental and psychiatric phenotypes.

### The observed enrichment architecture is not driven by individual loci or analytic choices

Given the modest effect sizes observed across individual phenotypes, we next evaluated whether the overall pattern of ND-CNV-associated enrichment remained stable across multiple orthogonal sensitivity analyses. A major strength of this study is the robustness of the observed signal across multiple analytic conditions. Associations remained stable following adjustment for healthcare utilization and ancestry-related covariates, exclusion of mCA-suspected variants, and leave-one-locus-out analyses, indicating that the observed enrichment pattern was not attributable to differential ascertainment, population structure, modeling choices, or a small number of influential loci. Permutation analyses provided complementary evidence that the aggregate phenotype architecture exceeded expectations under randomized exposure assignment.

Together, these findings support a model of distributed ND-CNV-associated neuropsychiatric liability spanning multiple recurrent loci and correlated phenotypes. This finding is consistent with models in which neuropsychiatric liability reflects contributions from multiple recurrent ND-CNV loci rather than a single dominant region. While certain loci (e.g., 22q11.2, 16p11.2) have well-characterized effects^24,37–40,55^, rare structural variant liability can reflect contributions from multiple regions and genes ^19,58^. Broad concordance between recurrent ND-CNV frequencies observed in All of Us and gnomAD-SV supports the interpretability of the overlap-based carrier framework. However, these comparisons are best viewed as population-frequency benchmarks rather than direct replication analyses due to differences in cohort composition and structural variant ascertainment. Thus, our results suggest that this distributed architecture is detectable even in a heterogeneous biobank setting, and that analytic frameworks focusing on single-locus effects may understate the broader structure of ND-CNV-associated risk.

### Correlated phenotypes may better capture ND-CNV-associated liability than burden counts

The domain-level analyses further support a model of distributed neuropsychiatric and transdiagnostic liability as a proxy for more sophisticated batteries of cognitive and behavioral data not yet available in the AoU^5,12,13,17^. Although no individual RDoC domain reached nominal significance, effect estimates were consistently shifted in a positive direction across multiple domains, particularly those plausibly linked to cognitive and affective function. At the same time, cross-domain burden metrics did not show strong separation between carriers and non-carriers. This apparent discrepancy is informative. The correlation analyses demonstrate that domains are moderately interdependent, while burden measures are highly correlated with one another. As a result, simple threshold-based burden metrics (e.g., ≥2 or ≥3 domains) may fail to capture the nuanced, distributed structure of ND-CNV effects. This finding is consistent with prior work suggesting that psychiatric and neurodevelopmental traits share substantial genetic architecture and are not well summarized by coarse aggregation metrics^27^. Instead, our results suggest that ND-CNV-associated risk appears to manifest as coordinated shifts across partially overlapping phenotypic dimensions, rather than uniform accumulation across domains.

### From recurrent loci to shared neuropsychiatric vulnerability

Taken together, these findings are consistent with mechanistic frameworks in which heterogeneous ND-CNV loci influence overlapping neurodevelopmental and neuropsychiatric processes involving synaptic function, circuit development, and broader gene regulatory networks ^10,26,52,56,59^. Although the present analyses do not directly evaluate molecular convergence, the distributed phenotypic architecture observed across recurrent ND-CNV loci is compatible with models in which diverse genomic perturbations influence partially shared biological systems, resulting in correlated effects across multiple neuropsychiatric domains (Fig. 8). Rather than supporting a one-to-one mapping between individual loci and discrete diagnoses, our results are more consistent with a model of distributed neuropsychiatric liability in which recurrent ND-CNVs contribute to overlapping dimensions of psychiatric and developmental vulnerability.

**Figure 8.**
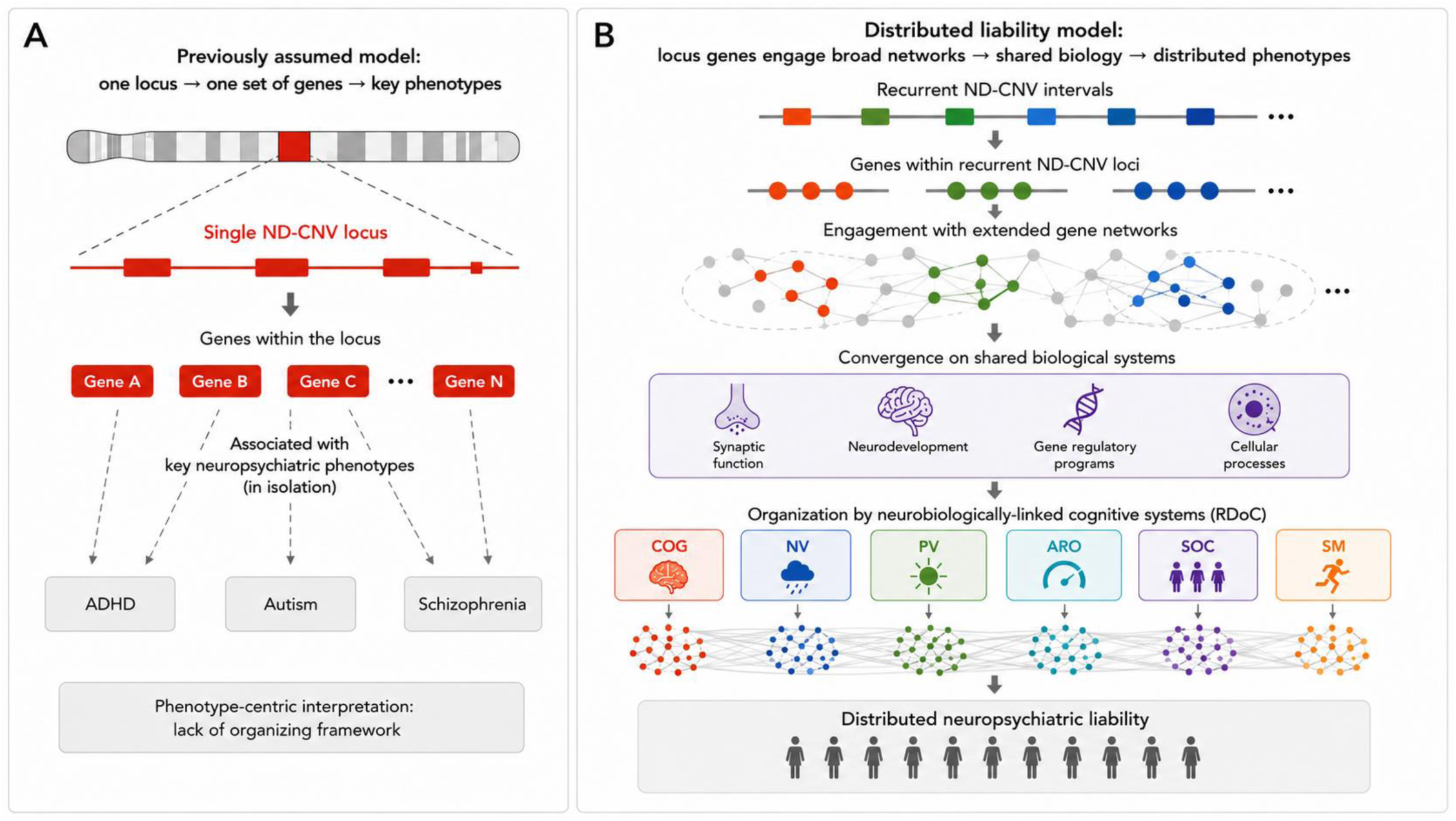
Conceptual framework for distributed neuropsychiatric liability associated with recurrent ND-CNVs. **(A)** Traditional phenotype-centric models have often interpreted recurrent neurodevelopmental copy number variants (ND-CNVs) as acting through dosage changes within a single locus to influence a limited set of discrete neuropsychiatric diagnoses. Under this framework, genes contained within an individual ND-CNV interval are viewed primarily in relation to specific downstream phenotypes, without a broader organizing structure linking phenotypic outcomes. **(B)** Conceptual model informed by the present findings. Genes within recurrent ND-CNV loci are proposed to engage broader biological networks extending beyond individual intervals, contributing to convergence on partially overlapping biological systems involved in synaptic function, neurodevelopment, gene regulatory programs, and cellular processes. Organizing phenotypes according to neurobiologically linked cognitive systems within the Research Domain Criteria (RDoC) framework reveals distributed patterns of neuropsychiatric liability spanning multiple correlated domains. In this interpretation, recurrent ND-CNVs are not expected to map onto single neuropsychiatric disorders in a one-to-one manner, but instead contribute to broad and correlated phenotypic architectures reflecting shared systems-level vulnerability.

### Interpreting distributed signal in the absence of phenotype-specific significance

Under conventional Benjamini–Hochberg FDR correction, no individual phenotype associations remained significant. However, the correlation structure among neuropsychiatric phenotypes and domains suggests that these tests are not independent, potentially reducing sensitivity to detect distributed effects spanning multiple related outcomes. Accordingly, the absence of FDR-significant associations should not be interpreted as evidence of no true signal, but rather as reflecting modest effect sizes distributed across correlated phenotypes, as interpreted by others^27,60,61^.

Consistent with this interpretation, global permutation analyses demonstrated significant aggregate directional enrichment for pooled, deletion-overlapping, and duplication-overlapping ND-CNV models despite the absence of individual FDR-significant associations. Together with the positive-control benchmarking, domain-level analyses, literature concordance, and robustness analyses, these findings suggest that the primary signal in this dataset reflects structured neuropsychiatric enrichment distributed across correlated phenotypes rather than a small number of large-effect phenotype-specific associations. This distributed architecture also has implications for replication. Validation of aggregate ND-CNV enrichment patterns will likely require large population-scale cohorts with harmonized structural variant callsets, longitudinal phenotyping, and sufficient sample sizes to capture rare recurrent ND-CNVs.

### Limitations and future directions

Several limitations should be considered when interpreting these findings. First, the present study employed a curated-locus, hypothesis-driven framework focused on recurrent ND-CNV regions with established neurodevelopmental, cognitive, or psychiatric relevance. While this approach improves interpretability and reduces multiple-testing burden, it was not designed to discover novel pathogenic CNVs. Structural variants outside the curated interval set, including rare, ancestry-specific, or incompletely characterized loci, were therefore not evaluated. Second, neuropsychiatric phenotypes were derived from electronic health records rather than standardized research assessments. Consequently, diagnostic coding reflects healthcare utilization, ascertainment, and documentation practices in addition to underlying neuropsychiatric liability, and absence of a diagnosis should not be interpreted as absence of disease.

Third, although global enrichment analyses demonstrated significant aggregate signal, no individual phenotype association survived false discovery rate correction. These findings therefore support distributed neuropsychiatric enrichment rather than definitive phenotype-specific associations. Future multivariate and latent-variable approaches may improve power to detect correlated effects across phenotypes. Finally, the overlap-based carrier framework introduces important analytical considerations. Although sensitivity analyses yielded broadly concordant results across alternative specifications, partially overlapping events may differ in breakpoint structure and biological consequences. Replication in additional large-scale genomic cohorts with harmonized structural variant callsets and longitudinal phenotyping will be important for evaluating the generalizability of these findings.

## CONCLUSION

In summary, this study demonstrates that ND-CNV carrier status in a large population biobank is associated with a distributed, pleiotropic pattern of neuropsychiatric risk that is robust across analytic conditions. Overall, these findings support the feasibility of detecting distributed ND-CNV-associated neuropsychiatric signal within large EHR-linked biobanks and are consistent with models of shared systems-level vulnerability across recurrent neurodevelopmental CNV loci. This framework provides a foundation for future efforts to integrate genetic, biological, and clinical data in the study of neurodevelopmental and psychiatric disorders. More broadly, these findings support the utility of large-scale EHR-linked genomic biobanks for studying distributed and pleiotropic neuropsychiatric risk architectures that may not be readily captured within traditional case-control design.

## METHODS

### Ethical Statement and Data Governance

Analyses were conducted using controlled-tier data from the All of Us Research Program (Controlled Data Release v8; CDRv8) within the Researcher Workbench under approved data use agreements. The All of Us Research Program has institutional review board (IRB) approval, and all participants provided informed consent for use of their de-identified data in research. Analyses adhered to program-specific privacy and reporting guidelines, including suppression of small cell counts and exclusion of participant identifiers from exported results. Certain locus-specific outputs were subject to All of Us privacy-policy enforcement thresholds and disclosure-protection procedures, including suppression of low-count or unstable locus-domain combinations in exported summary tables and visualization layers.

### Study population

Analyses were conducted using controlled-tier data from the All of Us Research Program (Controlled Data Release v8; CDRv8) within the Researcher Workbench. Participants were eligible if they had consented to use of electronic health record (EHR) and genomic data and had available short-read whole genome sequencing (WGS) data with corresponding structural variant (SV) calls.

Quality control (QC) and cohort definition were performed primarily using curated participant- and variant-level flags provided by the All of Us program. These included standardized indicators of sequencing quality, variant call confidence, and data completeness, allowing restriction to high-quality samples and minimizing reliance on study-specific filtering. Participants were required to pass all program-level QC filters for genomic data release, including inclusion in the high-quality WGS cohort and availability of derived genetic ancestry assignments and principal components.

Structural variant calls were obtained from the CDRv8 short-read SV callset and processed within a distributed Hail framework. Calls were harmonized into a unified representation including chromosome, genomic coordinates, SV type, and participant identifier. Analyses were restricted to autosomal deletions and duplications, and variants were filtered to retain high-confidence calls based on AoU-provided QC annotations, including call-level quality metrics and recommended inclusion flags. Variants failing these criteria or lacking complete annotation were excluded.

Additional preprocessing steps included removal of variants with implausible genomic coordinates or lengths, harmonization of contig naming to GRCh38 conventions, and restriction to biallelic representations where applicable. To reduce potential confounding from somatic events, variants consistent with mosaic chromosomal alterations (mCAs) were flagged using heuristic criteria derived from size, genomic span, and call characteristics, and were excluded in sensitivity analyses.

Participant-level inclusion further required sufficient EHR data for phenotype ascertainment and complete covariate information, including age, sex, genetic ancestry, and healthcare utilization measures. Analyses were restricted to unrelated individuals based on genome-wide kinship estimates, retaining a single representative from each related pair exceeding a predefined relatedness threshold.

Final analytic datasets were constructed using complete-case inclusion, with all QC steps applied prior to downstream aggregation of SV calls to curated interval sets. The resulting cohort reflects a high-confidence, population-based sample suitable for evaluating ND-CNV associations in a real-world biobank setting. All preprocessing and QC steps were implemented within a reproducible, modular pipeline using Hail for distributed genomic processing and pandas-based workflows for downstream aggregation, ensuring consistent application of filtering criteria across all analyses.

### Curated interval resources

Neurodevelopmental CNV (ND-CNV) loci were defined a priori using external literature and curated genomic resources to prioritize recurrent CNV regions with established neurodevelopmental, cognitive, or psychiatric relevance. This curated-locus strategy was selected to maximize biological interpretability and reduce multiple-testing burden in the primary analysis. CNVs outside these predefined intervals were not classified as ND-CNV exposures in the current study, even if they may have potential clinical or biological relevance. Therefore, the carrier definitions used here reflect known ND-CNV loci rather than an exhaustive genome-wide pathogenic CNV discovery framework.

Genomic coordinates were harmonized to GRCh38, and loci failing contiguous UCSD liftover were excluded. Final intervals were validated against external resources (e.g., ClinGen, OMIM), yielding a curated set of autosomal ND-CNV loci. Both raw and padded (±100 kb) interval sets were generated to evaluate sensitivity to boundary definition. Matched control intervals were constructed for each ND-CNV locus using feature-matching on chromosome, interval length, GC content, and segmental duplication burden. All intervals were represented as BED-formatted resources and ingested into the analytic pipeline.

### Structural variant data, preprocessing, and carrier definition

SV calls were obtained from chromosome-specific CDRv8 callsets and standardized into a unified long-format representation containing participant identifiers, genomic coordinates, SV type, and call-level metadata. Calls were restricted to high-confidence deletions and duplications overlapping curated interval regions. Each SV was annotated with interval overlap metrics, length, and genomic position, forming the basis for downstream carrier assignment. SVs were intersected with curated ND-CNV and matched control intervals using interval-based operations. For each SV–interval pair, multiple overlap metrics were computed, including base-pair overlap, fractional overlap relative to both SV and interval, and midpoint inclusion.

Carrier status was defined using rule-based overlap criteria, with the primary definition based on reciprocal overlap ≥30% (“ro30”). Stricter thresholds (ro50, ro70) and alternative definitions were evaluated in sensitivity analyses. Participant-level carrier status was assigned if at least one SV satisfied the primary criterion for any ND-CNV locus. Additional exposure variables captured deletion-overlapping and duplication-overlapping carrier status, as well as the assigned count of curated ND-CNV loci per participant.

To evaluate the effect of interval-overlap stringency on recurrent ND-CNV assignment, observed structural variant calls from the All of Us srWGS resource were intersected against curated recurrent ND-CNV intervals using multiple overlap definitions, including reciprocal-overlap thresholds (ro30, ro50, ro70) and fractional locus-overlap criteria. Participant-level carrier counts were summarized across overlap frameworks to assess the balance between sensitivity and interval specificity. External benchmarking analyses compared recurrent ND-CNV intervals against the gnomAD-SV v2.1 population structural variant resource using direction-matched reciprocal-overlap definitions (ro30 and ro50) and non-neuro subset allele-frequency annotations. These analyses were intended as contextual population-frequency benchmarking rather than formal replication analyses.

### Matched negative interval control framework

To assess specificity, matched genomic control intervals were constructed for each ND-CNV locus using feature-matching on chromosome, interval length, GC content, and segmental duplication burden. SV overlap and exposure assignment were performed identically for ND-CNV and control intervals, enabling direct comparison of signal against a matched genomic background. Control intervals were selected from genomic regions not included in the curated ND-CNV locus set and lacking established neurodevelopmental pathogenicity annotations.

### EHR phenotype ascertainment, case definitions, and limitations

Neuropsychiatric phenotypes were ascertained from electronic health record (EHR) data using OMOP condition concept mappings. For each phenotype, participant-level case status was defined as the presence of ≥1 qualifying diagnosis code within the observation window. Controls were defined as participants without any recorded diagnosis corresponding to the phenotype of interest. Phenotypes were treated as prevalent conditions, reflecting lifetime diagnostic history captured in the EHR rather than incident onset. To ensure stability of model estimates, phenotypes with insufficient case counts were excluded from downstream analyses. All phenotypes were constructed using standardized concept hierarchies, including descendant terms, to harmonize across coding systems and maximize sensitivity of case detection. The full phenotype definitions and mappings are provided in Supplemental Table S3.

Several limitations related to EHR-based ascertainment should be considered when interpreting these findings. Psychiatric and neurodevelopmental phenotypes within All of Us reflect observed diagnostic coding patterns rather than standardized research assessments, and absence of a recorded diagnosis should not be interpreted as definitive absence of disease. Accordingly, these analyses are best interpreted as evaluating EHR-observed phenotypic burden and directional enrichment patterns within a large population biobank framework rather than definitive syndrome prevalence or fully penetrant clinical expression.

### Statistical Analysis

Associations between ND-CNV carrier status and neuropsychiatric phenotypes were evaluated using logistic regression with maximum likelihood estimation. All primary logistic regression models were adjusted for age at last observation, log-transformed healthcare utilization (log1p number of unique clinical visits), observation time (days of EHR follow-up), and the first five genetic ancestry principal components (PC1–PC5). Separate models were constructed for pooled ND-CNV exposure, deletion-overlapping and duplication-overlapping ND-CNV carriers, matched control intervals, and alternative overlap definitions. All logistic regression models were evaluated for convergence and numerical stability. Models with insufficient case counts (N ≤ 20) or evidence of complete or quasi-complete separation were excluded from analysis. Continuous covariates were inspected for appropriate scaling, and model outputs were assessed for implausible effect estimates or inflated confidence intervals. These steps ensured robustness of estimated associations across phenotypes and analytical conditions.

Results are reported as odds ratios (ORs) with 95% confidence intervals (CIs). To characterize structured pleiotropy, analyses were extended to the domain level by aggregating phenotypes within RDoC categories. Cross-domain burden measures were defined based on the number of affected domains per individual. To account for multiple hypothesis testing, p-values were adjusted using the Benjamini–Hochberg false discovery rate (FDR) procedure. Correction was applied separately within predefined model families, including phenotype-level analyses, domain-level analyses, and sensitivity analyses, to preserve interpretability within each analytical context. Both nominal (p < 0.05) and FDR-adjusted significance thresholds are reported. Aggregate enrichment metrics derived from phenotype-level effect estimates were additionally used to compare sensitivity analyses and quantify changes in overall enrichment architecture following locus exclusion.

### Global permutation enrichment analysis

To evaluate whether the observed pattern of ND-CNV-associated neuropsychiatric effects exceeded that expected under random exposure assignment, we performed a phenotype-level exposure-permutation analysis. Using the participant-level modeling dataset generated for the primary analyses, ND-CNV exposure labels were randomly reassigned across participants while preserving the observed phenotype structure, phenotype prevalences, and analytic framework. For each permuted dataset, the full set of phenotype-level logistic regression models was re-estimated using the same covariate specification and modeling procedures applied in the primary analyses.

Global directional enrichment was quantified using the summed phenotype-level Wald Z-statistic (ΣZ), which preserves both effect direction and magnitude across all modeled neuropsychiatric phenotypes. A total of 1,000 permutations were performed for pooled ND-CNV carrier status, deletion-overlapping carrier status, and duplication-overlapping carrier status. Empirical permutation probabilities were calculated by comparing the observed ΣZ statistic with the corresponding permutation-derived null distribution. Secondary summary metrics, including the proportion of phenotypes with positive effect estimates, were evaluated to assess directional consistency across phenotype-level associations.

### Exposure Scaling and Effect Interpretation

Primary analyses modeled ND-CNV carrier status (ro30) as a binary exposure. Direction-specific analyses further distinguished deletion-overlapping and duplication-overlapping ND-CNV carrier assignments. Because participants could contribute to both exposure groups, these analyses should be interpreted as non-mutually-exclusive exposure models unless otherwise specified. For ND-CNV locus-count analyses, exposure was defined as the number of curated ND-CNV loci assigned to each participant under the overlap-based carrier framework and modeled as a continuous variable. Domain-level phenotypes and cross-domain burden measures were similarly represented as binary or count-based variables, depending on the analysis. Effect estimates are therefore interpreted as the relative odds of phenotype presence per exposure definition.

### Covariates

All regression models were adjusted for factors expected to influence diagnostic ascertainment, follow-up opportunity, and population structure within the All of Us cohort. Covariates included age at last observation, healthcare utilization (log-transformed number of unique clinical visits), observation time (days of EHR follow-up), and the first five genetic ancestry principal components (PC1–PC5). Healthcare-utilization and observation-time covariates were included to account for differences in diagnostic opportunity across participants, while ancestry principal components were included to mitigate potential confounding due to population structure. The same covariate framework was applied across primary phenotype-level analyses, domain-level analyses, positive-control benchmarking, and sensitivity analyses unless otherwise specified.

### Sensitivity and robustness analyses

Robustness of observed associations was evaluated through a series of complementary sensitivity analyses designed to assess the influence of locus definition, variant composition, and model specification. Alternative overlap thresholds (reciprocal overlap ≥50% and ≥70%) were examined to evaluate sensitivity to carrier definition, alongside matched genomic control intervals to assess specificity relative to background structural variation. Direction-specific analyses compared deletion-only and duplication-only carriers to assess differential effects by variant type. To address potential confounding from somatic variation, analyses were repeated after exclusion of variants suspected to represent mosaic chromosomal alterations (mCAs).

Leave-one-locus-out (LOLO) analyses were performed to quantify the contribution of individual loci to association signals and to evaluate whether observed effects were driven by specific regions or reflected distributed genetic architecture. For each recurrent ND-CNV interval, all participants assigned to that interval under the primary ro30 framework were excluded and the aggregate enrichment metrics were recalculated. Deviations from the full ND-CNV model were used to estimate locus-specific influence on the overall enrichment architecture. To facilitate interpretation of influential loci identified through LOLO analyses, locus-specific phenotype association profiles were generated using the primary ro30 exposure framework. For visualization, nominally significant phenotype associations (P < 0.05) were summarized across influential recurrent loci.

Finally, ancestry-stratified analyses were conducted to assess consistency of associations across population subgroups. Across these analyses, effect estimates were compared for stability in direction and magnitude, providing a comprehensive assessment of robustness to ascertainment, locus composition, and modeling assumptions.

To contextualize observed ND-CNV phenotype enrichment patterns within the broader neurodevelopmental CNV literature, we performed a qualitative domain-level concordance analysis comparing recurrent locus associations reported in prior studies with directional enrichment patterns observed in All of Us. Canonical ND-CNV loci and associated phenotype domains were manually curated from prior CNV meta-analyses, registry cohorts, population-biobank studies, and cross-disorder neurodevelopmental investigations^13,17,53,62–69^. Literature-supported phenotype-domain relationships were assigned qualitative evidence scores and compared against AoU-derived locus-specific directional enrichment patterns generated from ro30 participant-level exposure models. This analysis was intended as a contextual and hypothesis-generating comparison rather than a formal meta-analysis.

### Computational environment and reproducibility

All analyses were conducted within the All of Us Researcher Workbench using a Google Cloud Dataproc environment with Apache Spark and Hail for distributed genomic processing. SV extraction, interval overlap, and participant-level aggregation were implemented in Hail, while downstream statistical analyses were performed in Python using pandas and statsmodels. The analytic workflow was implemented as a modular, reproducible pipeline with explicit intermediate datasets corresponding to each processing stage. Final analytic datasets, model outputs, and summary tables are provided as structured supplemental resources, with additional pipeline details described in the supplemental methods. Extended methods are available in the Supplemental Materials. Data provenance summaries for main and supplemental figures, along with source code and associated analysis outputs, are provided to support reproducibility.

## Data Availability

Full pipeline outputs for this project have been deposited into Open Science Framework.

https://osf.io/5dxtv/overview?view_only=4a86da9f04c6456d871ca9cf96e9027b

