## Supplement for "Pleiotropic and Distributed Neuropsychiatric Effects of Neurodevelopmental Copy Number Variants in the All of Us Biobank"

**Short Title: A Scalable Framework for ND-CNV Risk in *All of Us***

Anne Marie Wells, PhD[1-3], Feiyang Zhao, PhD[2], Sudha Seshadri, MD[2], Jose Cavazos, MD, PhD[2,3], Agustin Ruiz, MD, PhD[3-6]

**Author Affiliations**
1. South Texas Medical Scientist Training Program, UT Health San Antonio, San Antonio, TX
2. Glenn Biggs Institute for Alzheimer's and Neurodegenerative Diseases, UT Health San Antonio, San Antonio, TX
3. Long School of Medicine, UT Health San Antonio, San Antonio, TX
4. Department of Microbiology, Immunology and Molecular Genetics. Long School of Medicine, The University of Texas at San Antonio, San Antonio, TX, USA
5. Ace Alzheimer Center Barcelona-Universitat Internacional de Catalunya, Barcelona, Spain
6. CIBERNED, Network Center for Biomedical Research in Neurodegenerative Diseases, National Institute of Health Carlos III, Madrid, Spain

**Corresponding Author**
Anne Marie Wells, PhD


**Table of Contents**


| Section | Title | Page |
|---|---|---|
| I | External interval resource curation | 2 |
| II | Phenotype construction and dictionary mapping | 5 |
| III | Extended Pipeline Methods | 7 |
| V | Supplemental Figures | 18 |
| VI | Supplemental Tables | 30 |
| VI | All of Us (AoU) and AI Disclosures | 31 |

### I. External interval resource curation

#### Curation of neurodevelopmental CNV (ND-CNV) intervals

A curated set of neurodevelopmental copy number variant (ND-CNV) loci was assembled through systematic aggregation of CNV call lists from large-scale studies demonstrating significant associations with neurodevelopmental and psychiatric phenotypes, including Thygesen et al. 2021, Stefansson et al. 2014, and Kendall et al. 2017. These sources were used to construct an initial exhaustive list of candidate loci (**Supplement Table 3**).

Because reported interval coordinates varied across genome reference builds, all loci were harmonized to the GRCh38 reference genome using the UCSC LiftOver tool. Intervals that failed to map contiguously to GRCh38 (e.g., those resulting in split or multi-segment mappings) were excluded to ensure consistent and interpretable locus definitions. Following coordinate harmonization, ND-CNV intervals were cross-referenced against major genomic databases of clinically and biologically validated loci, including ClinGen, ClinVar, DECIPHER, and OMIM, to confirm concordance with established consensus regions.

Consistent with prior literature, and reflecting the biology of recurrent CNV loci, reciprocal deletions and duplications were treated as sharing identical genomic boundaries at most loci. Accordingly, interval definitions were harmonized such that deletion and duplication representations corresponded to the same genomic coordinates unless strong evidence supported asymmetric boundaries.

To support sensitivity analyses of structural variant (SV) detection and overlap definitions, a secondary padded interval set was generated by extending each locus symmetrically by ±100 kb. Final curated intervals were exported as BED files in GRCh38 coordinates, comprising 96 total ND-CNV loci:

- nd_cnv_loci_GRCh38_pad0kb_v1.0.bed.bed (raw)
- nd_cnv_loci_GRCh38_pad100kb_v1.0.bed.bed (padded)

Both raw and padded interval sets were retained and propagated through downstream analyses. The final ND-CNV interval resource was exported as tabular files and ingested into Hail tables (ht_nd_raw, ht_nd_padded) for integration into the analytic pipeline.

#### Deduplication and preparation of loci for negative control interval generation

For the purpose of generating matched control intervals, ND-CNV loci were collapsed to a non-redundant set of genomic intervals. This deduplication step accounted for loci represented multiple times across source studies or across reciprocal CNV types (deletion and duplication), resulting in a reduced set of 51 unique genomic intervals:

- ND_CNV_loci_hg38_intervalmatch_raw.bed (raw)
- ND_CNV_loci_hg38_intervalmatch_padded_100kb.bed (raw)

These deduplicated loci were used as the reference set for control interval matching, ensuring that control interval generation reflected the underlying genomic distribution of unique ND-CNV regions rather than duplicated representations of the same locus.
To support robustness analyses of overlap definitions, padded versions of each ND-CNV interval were generated by symmetrically extending canonical boundaries by a fixed window (100 kb in sensitivity analyses).

#### Generation of matched negative control intervals

Matched genomic negative control intervals were generated for each neurodevelopmental CNV locus using a custom local pipeline implemented in Jupyter Notebook within an Ubuntu environment running under Windows Subsystem for Linux. Input loci were provided as hg38 BED intervals representing collapsed ND-CNV regions, such that each recurrent deletion or duplication locus was represented by a single interval prior to control generation. Intervals were loaded into Python, assigned standard genomic interval fields (chrom, start, end, and, where available, name), and annotated with interval length calculated as end - start.

GC content for each ND-CNV interval was calculated using bedtools nuc against the hg38 reference genome. Segmental duplication burden was estimated using the UCSC hg38 genomicSuperDups annotation, downloaded from the UCSC Genome Browser database resource. The relevant genomic coordinates were converted to BED format, and ND-CNV intervals were intersected with the segmental duplication intervals using bedtools intersect -wao. For each locus, the total number of overlapping base pairs was summed and divided by interval length to obtain the segmental duplication fraction. Chromosome sizes were obtained from an hg38 chromosome-size file and restricted to chromosomes represented in the ND-CNV set.

For each ND-CNV locus, 1,000 candidate control intervals were generated on the same chromosome and with the exact same length as the target locus. Candidate start positions were sampled uniformly such that the resulting interval remained fully within chromosome boundaries. GC content for candidate intervals was computed from the hg38 FASTA reference using indexed sequence retrieval with pysam, and segmental duplication fraction was computed using the same segmental duplication annotation used for the ND-CNV loci. Candidate intervals were then filtered to retain only those within an absolute GC-content difference of 0.02 and an absolute segmental-duplication-fraction difference of 0.05 relative to the corresponding ND-CNV locus. For each locus, all passing candidates were retained if 10 or fewer intervals met these criteria; if more than 10 candidate intervals passed filtering, 10 were selected by random sampling with a fixed random seed (random_state=42). Thus, each locus contributed between 0 and 10 matched control intervals, depending on the number of candidates satisfying the prespecified matching thresholds.

As a quality-control step, we compared the distributions of interval length, GC content, and segmental duplication fraction between ND-CNV loci and matched control intervals by plotting feature-wise density histograms. The final matched control interval set was then exported as a tab-delimited file.

**Sensitivity analysis using 100 kb-padded loci**

To evaluate sensitivity to locus definition, the matching procedure was repeated using a second set of ND-CNV intervals padded by 100 kb. These padded loci were re-annotated for GC content and segmental duplication fraction and subjected to the same control-generation workflow used in the primary analysis. Specifically, 1,000 same-chromosome, same-length candidate intervals were generated per padded locus; candidate intervals were filtered using the same absolute tolerances for GC content and segmental duplication fraction; and up to 10 passing intervals were retained per locus, with random downsampling to 10 when more than 10 candidates satisfied the filtering criteria. The resulting sensitivity-analysis control set was exported separately.

**Software and computational environment**

Analyses were run locally under Ubuntu in Windows Subsystem for Linux. System-level tools installed for the workflow included bedtools, wget, gzip, and python3-venv. A dedicated Miniconda environment (cnv_env) was created with Python 3.11, and notebook-based analyses used at least jupyterlab, pandas, numpy, tqdm, and pysam.

**Data and code availability**

Control interval generation was implemented in standalone preprocessing pipelines executed outside the main analytic notebook. These pipelines included:

- A local run script for initial control interval matching and filtering (neg controls local run notebook)
- A sensitivity-analysis pipeline for generating padded control intervals and evaluating robustness to interval expansion (neg controls_sensitivity_analysis_padded100kb notebook)

Both pipelines operated on tabular genomic interval representations and produced final matched control interval sets exported as TSV files. Intermediate steps included computation of genomic feature annotations, candidate interval generation, feature-based matching, and validation of matching quality. The final control interval resources were versioned and treated as fixed inputs to the main analytic pipeline. Upon ingestion, control intervals were converted to Hail tables (ht_ctrl_raw, ht_ctrl_padded) and harmonized with ND-CNV interval resources into a unified interval framework (ht_intervals_all).

The code used to generate matched negative control intervals and the corresponding 100 kb-padded sensitivity-analysis control set will be released as an external package upon publication. The release will include the interval-generation workflow, dependency specifications, and documentation describing required reference inputs, including the hg38 reference genome and segmental duplication annotations. Processed matched-control interval files generated for this study are available in **Supplement Table 3**, as well as external package files:

- nd_matched_controls.segdup_gc_matched.tsv (raw)
- nd_padded_matched_controls.segdup_gc_matched.tsv (padded)

Each interval was annotated using the same schema as ND-CNV loci, including interval identifiers, matching features (length, GC content, segmental duplication fraction), and CNV-type placeholders for compatibility with downstream analyses.

**Integration with downstream analyses**
The curated ND-CNV and matched control interval resources define the genomic search space for all downstream overlap and exposure-mapping steps. Observed structural variant calls were intersected with both ND-CNV and control interval sets using identical overlap criteria and rule-based assignment logic. This parallel treatment enables direct comparison of ND-CNV and control interval exposures, supporting evaluation of the specificity of observed associations and providing an internal negative control framework for the analytic pipeline.

### II. Phenotype construction and dictionary mapping

#### Phenotype dictionary construction

Psychiatric phenotypes were defined using a structured, dictionary-driven framework integrating All of Us (AoU) OMOP condition concepts with a domain-based classification scheme aligned to Research Domain Criteria (RDoC) constructs and clinically relevant diagnostic groupings. This approach was designed to support both phenotype-specific analyses and higher-order representations of structured pleiotropy across neuropsychiatric domains.

A custom phenotype dictionary was developed to map OMOP condition concepts to psychiatric phenotypes of interest. The dictionary was constructed by integrating AoU-native OMOP concept identifiers corresponding to psychiatric diagnoses with hierarchical expansion to descendant concepts using OMOP concept relationships. Each phenotype definition was annotated with both dimensional and categorical metadata, including a primary domain assignment aligned to RDoC constructs, an optional secondary domain assignment capturing cross-domain clinical features, and a group-level label reflecting DSM-style diagnostic groupings. Primary domain assignments were determined based on the dominant clinical features of each phenotype, while secondary domain assignments were used to capture known overlap in symptom dimensions across domains.

Given the multidimensional nature of psychiatric phenotypes, domain assignments were designed to be non-mutually exclusive, allowing individual phenotypes to contribute to multiple domains where clinically appropriate. This hybrid mapping approach balances dimensional (RDoC-aligned) and categorical (DSM-informed) representations of psychiatric phenotypes, enabling analyses that capture both structured symptom domains and clinically interpretable diagnostic groupings.

#### OMOP concept expansion and condition extraction

Phenotype definitions were operationalized by expanding each dictionary-defined OMOP concept to include all descendant concepts within the OMOP hierarchy, ensuring comprehensive capture of clinically related diagnoses across coding systems and levels of specificity. Condition occurrence data were extracted from the AoU database using SQL queries restricted to these expanded concept sets. Extracted records were stored in a long-format table and subsequently annotated with phenotype dictionary metadata, linking each condition occurrence to its corresponding phenotype label, domain assignments, and group-level classification.

#### Participant-level phenotype flag generation

Condition-level data were collapsed to participant-level binary phenotype indicators by grouping records by participant identifier and phenotype. Individuals were classified as cases for a given phenotype if they had at least one recorded condition occurrence corresponding to any concept within the expanded concept set. Participants without any recorded diagnosis for a given phenotype were classified as controls for that phenotype, conditional on inclusion in the analytic cohort. This process yielded a participant-level phenotype matrix containing one row per individual and binary indicators for all phenotypes of interest.

#### Domain-level and grouped phenotype representations

To support analyses of structured pleiotropy, phenotype-level indicators were further aggregated into domain-level and group-level representations. Domain-level indicators were constructed by collapsing phenotype flags within each primary domain category, such that individuals were classified as domain-positive if they had at least one phenotype assigned to that domain. Secondary domain assignments were retained in these aggregations, allowing phenotypes to contribute to multiple domains and preserving cross-domain structure. Group-level indicators were constructed by aggregating phenotypes within clinically defined groupings, enabling complementary analyses aligned with DSM-style diagnostic categories.

#### Integration with analytic pipeline

Participant-level phenotype flags and derived domain and group indicators were merged with demographic, genomic, and exposure variables to form the canonical analytic dataset. These phenotype variables were propagated through all downstream analyses, including primary ND-CNV exposure models, matched control comparisons, domain-level analyses, and multi-domain burden models. Phenotype labels and domain assignments were further used to annotate regression outputs and visualizations, ensuring consistent interpretation across descriptive summaries, statistical models, and figures.

A complete mapping of phenotype definitions, OMOP concept identifiers, domain assignments, and group-level classifications is provided in **Supplemental Table 3**.

### III. Extended Pipeline Methods

#### Study design and analytic environment

All analyses were performed in the All of Us Researcher Legacy Workbench using a Hail- and Python-based pipeline implemented in a single structured notebook. Genomic and phenotypic data were accessed through the All of Us Researcher Workbench using the Controlled Tier dataset (CDRv8; release C2024Q3R4, February 3, 2025). Analyses were conducted using a Google Cloud Dataproc cluster configured for large-scale genomic data processing. The analytic environment leveraged Apache Spark via Hail for distributed computation over chromosome-specific MatrixTables and interval resources. The Dataproc cluster was provisioned with scalable worker nodes to support parallelized processing of structural variant extraction, interval overlap operations, and participant-level aggregation steps. Storage and intermediate datasets were managed using Google Cloud Storage, with data persisted in Hail-native and columnar formats to optimize I/O performance. Downstream statistical analyses were performed in Python (v3.10), PySpark (v3.5.3) using pandas, statsmodels, NumPy, matplotlib, and seaborn within the same cloud-based environment, enabling seamless transition between distributed and local data representations. This configuration supported efficient execution of end-to-end analyses across hundreds of thousands of participants and genome-wide interval datasets. Internal storage paths and workspace-specific file structures are not reported, as they are environment-dependent; however, all data transformations, derived variables, and analytic datasets are described in detail to enable reproducibility.

The workflow was organized into sequential modules for curated interval import and harmonization, cohort construction, extraction of observed structural variant (SV) calls, curated interval overlap assignment, participant-level exposure derivation, descriptive summarization, regression modeling, direction-specific analyses, figure generation, and export packaging. Primary data modalities included curated interval resources, chromosome-specific SV MatrixTables, participant-level demographic and genomic quality control data, ancestry predictions and principal components, and dictionary-driven electronic health record phenotype definitions. The analytic pipeline explicitly separated Hail-native large-scale operations from downstream pandas- and statsmodels-based modeling steps. Hail was used for interval handling, SV extraction, overlap assignment, participant-level aggregation, and construction of the analytic exposure table. Pandas was used for final model-ready table assembly and descriptive summaries. Logistic regression analyses were performed with statsmodels.api.Logit.

#### Structural variant preprocessing and chromosome-specific MatrixTable generation

Short-read whole-genome sequencing (srWGS) structural variant (SV) resources from the All of Us Research Program Controlled Tier (CDRv8) were first converted into chromosome-specific Hail MatrixTables as a prerequisite preprocessing step for downstream ND-CNV analyses. Because the native srWGS SV resources were distributed across large chromosome-partitioned VCF-derived datasets, preprocessing was performed to enable scalable interval-based overlap assignment, participant-level aggregation, and phenotype association analyses within the Hail/Spark framework.

For each autosome, SV calls were imported into Hail and harmonized to GRCh38 genomic coordinates. Structural variant metadata, including SV type (deletion or duplication), genomic interval boundaries, participant identifiers, and call-level annotations, were retained in chromosome-specific MatrixTables to facilitate efficient interval filtering and overlap operations. These chromosome-level resources were subsequently used to generate long-format SV call tables and overlap-assignment tables used throughout the ND-CNV analytic pipeline.

Following preprocessing, recurrent neurodevelopmental CNV (ND-CNV) intervals were intersected against observed SV calls using multiple interval-overlap definitions, including permissive locus-fraction criteria and reciprocal-overlap thresholds. Resulting overlap-assignment tables were then collapsed to participant-level exposure representations for downstream burden estimation, phenome-wide association analyses and robustness/sensitivity analyses.

The preprocessing and MatrixTable generation steps were performed entirely within the All of Us Researcher Workbench using Dataproc-backed Hail (ref: GRCh38) and Apache Spark cluster, and served as foundational infrastructure for all subsequent SV-based analyses.

**Curated ND-CNV and matched-control interval resources**

Curated interval resources were imported in Hail from BED files representing neurodevelopmental CNV (ND-CNV) loci and matched control intervals. Four primary interval tables were created: ht_nd_raw, ht_nd_padded, ht_ctrl_raw, and ht_ctrl_padded. Each table was harmonized to a common schema including chromosome, interval set, CNV type, locus identifiers, interval identifiers, length, GC content, segmental duplication fraction, and a Hail interval object. A combined harmonized interval table, ht_intervals_all, was then created by selecting common fields across interval families and unioning the component tables. A separate collapsed unique-locus interval resource, ht_nd_unique_raw, was also imported for support analyses focused on unique ND loci.

Interval harmonization relied on helper functions defined in the notebook, including clean_chrom, make_bed_interval, parse_cnv_type_from_name, parse_target_locus_name, and autosome_filter. Interval-family audits were generated to summarize row counts, target locus counts, match-group counts, and mean interval length.

**Cohort construction, demographics, EHR phenotypes, and genomic QC**

Participant-level cohort construction began with SQL extraction of person-level demographic information into person_df. A phenotype dictionary was then loaded into phenotype_dict_df and expanded to descendant OMOP concepts using SQL and post-query merging, yielding phenotype_concepts_df and phenotype_screen_df. Long-format condition_occurrence rows corresponding to dictionary-derived concepts were extracted into condition_df and joined back to dictionary metadata to create condition_annotated_df.

Observation-period summaries and healthcare-utilization summaries were retrieved separately as observation_df and utilization_df. Long-format condition data were collapsed to one row per participant in person_phenotype_df, producing participant-level binary psychiatric phenotype flags. Genomic QC metrics and flagged-sample resources were imported from AoU-controlled assets and merged into genomic_metrics_df. Sex concordance was classified using the helper classify_sex_concordance, and genomic inclusion filtering yielded analysis_cohort_df. Relatedness pruning was then applied using a remove-list imported from Hail, generating analysis_cohort_pruned_df.

Ancestry predictions and genotype principal components were joined in two stages. First, categorical ancestry information was added; second, principal components were parsed and expanded using parse_pca_features. The final canonical participant-level analytic table was stored as analysis_samples_df. For Hail-native downstream joins, this participant table was converted back to a keyed Hail table, ht_samples, after datetime fields were converted to string to avoid schema-conversion errors.

**Extraction of observed structural variant calls from chromosome MatrixTables**

Observed structural variant calls were extracted from chromosome-specific ND-CNV MatrixTables using the helper load_and_standardize_nd_cnv_mt, which standardized row, column, and entry field interpretation across resources. Additional helpers, including row_overlaps_any_curated_interval_expr and entry_is_called_expr, were used to identify called SV entries and restrict attention to calls overlapping curated interval payloads.

A per-chromosome interval payload was first constructed as interval_payload_df and split into intervals_by_chrom. Compact chromosome-level call tables were then extracted and unioned to produce a long-format SV call table, ht_sv_calls_long. Each observed call was assigned a stable observed_call_id and audited for structural consistency. Standardized call-level fields included participant identifier, chromosome, start and end position, SV type, deletion and duplication indicators, SV length, and interval representation.

**SV–curated interval overlap assignment and rule-based carrier evidence**
Before overlap assignment, the observed call table was cleaned and curated intervals were restricted to the primary analysis families in ht_intervals_main. Overlap operations were performed in Hail using compact interval payloads and a set of explicit helper functions: bp_overlap_expr, frac_overlap_call_expr, frac_overlap_interval_expr, frac_overlap_smaller_expr, call_midpoint_expr, and midpoint_in_interval_expr.

Potential overlaps between observed SV calls and curated intervals were assembled in ht_overlap_candidates. For each candidate pair, the pipeline computed base-pair overlap, fraction of the call overlapped, fraction of the curated interval overlapped, fraction of the smaller interval overlapped, and whether the call midpoint fell within the curated interval. These metrics were then translated into explicit rule flags in ht_overlap_candidates, including any_overlap, ro30, ro50, ro70, frac_locus_30, frac_smaller_50, and midpoint.

ND and control interval overlap evidence were collapsed separately. ND overlaps were collapsed directly to ht_overlap_collapsed_nd with participant-, interval-, and locus-level aggregation of rule support, deletion/duplication status, supporting-row counts, and maximum overlap metrics. Control intervals were collapsed using a two-stage sharded strategy to accommodate scale, yielding ht_overlap_collapsed_ctrl_harmonized. Harmonized ND and control overlap tables were then unioned into ht_overlap_collapsed. A passed_rules set was built for each retained participant-by-interval assignment, and the lean Section D to Section E handoff table was stored as ht_overlap_for_exposure.

Under the notebook's primary analysis framing, CNV exposure at curated ND loci was treated as direction-agnostic at the locus-presence level, with deletion and duplication burden retained as separate derived indicators for secondary analyses.

**Participant-level exposure table construction**
The Section D handoff table was first annotated with interval-family flags (is_nd, is_ctrl) and explicit boolean indicators for retained overlap rules (has_rule_ro30, has_rule_ro50, has_rule_ro70, and has_rule_frac_locus_30). After defensive de-duplication to one row per person_id × interval_set × interval_id, overlap evidence was collapsed to participant level in ht_participant_exposure.

The participant-level aggregation generated the primary binary exposures nd_any and ctrl_any, both defined under the ro30 overlap rule, together with ND sensitivity exposures nd_any_ro50, nd_any_ro70, and nd_any_frac_locus_30. Direction-specific burden indicators were also derived: nd_del_any, nd_dup_any, ctrl_del_any, and ctrl_dup_any. Count variables summarizing the number of implicated intervals or loci per participant were generated in parallel, including nd_n_loci_ro30, ctrl_n_intervals_ro30, nd_n_loci_ro50, nd_n_loci_ro70, and nd_n_loci_frac_locus_30. Participant-level exposure summaries were stored in ht_participant_exposure_summary.

This exposure table was then left-joined back to the full canonical cohort (ht_analysis_cohort) to create ht_analysis_cohort_with_exposure. Missing exposure values among participants without retained overlap evidence were deterministically filled as unexposed, thereby preserving the full analytic denominator. The final pandas analytic table used for downstream descriptive and regression analyses was exported as df_analysis.

**Hierarchical ND-CNV carrier mapping framework**
ND-CNV carrier status was defined through a rule-based hierarchical mapping from observed SV calls to curated locus-level and participant-level exposure indicators. Let $i$ index participants, $c$ index observed structural variant calls, and $\ell$ index curated ND-CNV loci or matched control intervals. Observed SV calls were extracted from chromosome-specific MatrixTables and represented in long format as ht_sv_calls_long. Curated intervals were represented in harmonized interval tables and restricted to primary interval families for overlap analyses. For each observed call–interval pair, overlap metrics were computed:

- base-pair overlap,
- fraction of the observed call overlapped,

- fraction of the curated interval overlapped,
- fraction of the smaller interval overlapped, and
- whether the observed call midpoint fell within the curated interval.

These metrics were converted into explicit rule indicators:
- ro30,
- ro50,
- ro70,
- frac_locus_30,
- frac_smaller_50,
- midpoint, and
- any_overlap.

For the primary analysis, participant-level ND-CNV exposure was defined under the ro30 rule. Thus, participant $i$ was classified as ND-exposed if any retained observed call overlapped any curated ND interval with ro30 support:

$$\text{nd\_any}_i = 1 \iff \exists(c,\ell)\text{:participant } i \text{ has an ND overlap assignment with rule ro30.}$$

Analogously, matched-control exposure was defined as:

$$\text{ctrl\_any}_i = 1 \iff \exists(c,\ell)\text{:participant } i \text{ has a control-interval overlap assignment with rule ro30.}$$

Sensitivity definitions replaced the primary ro30 criterion with stricter rule sets, generating nd_any_ro50, nd_any_ro70, and nd_any_frac_locus_30. Direction-specific burden indicators were derived by intersecting the primary ro30 exposure rule with call direction:
- nd_del_any: any ND-overlapping deletion,
- nd_dup_any: any ND-overlapping duplication,
- ctrl_del_any: any control-overlapping deletion,
- ctrl_dup_any: any control-overlapping duplication.

Participant-level count measures summarized the number of ND loci or control intervals implicated under each retained rule. This framework allows a single observed-call extraction pipeline to support primary carrier analyses, rule-sensitivity analyses, and deletion-versus-duplication contrasts without redefining the underlying overlap engine.

**Descriptive summaries**
Descriptive analyses were generated from df_analysis. Table F1 (table_f1) summarized total cohort size and prevalence of each exposure variable, including the primary ND and control exposures, sensitivity-rule exposures, and pooled deletion/duplication indicators. Table F2 (table_f2) summarized count distributions among exposed participants, reporting mean, standard deviation, median, first quartile, and third quartile for ND locus counts and control interval counts under each retained rule.

Phenotype prevalence summaries were generated in three forms. Table F3A (table_f3a) compared phenotype prevalence between ND-exposed and ND-unexposed participants across the retained ND overlap rules specified in ND_RULE_EXPOSURES. Table F3B (table_f3b) compared phenotype prevalence between ND-exposed and matched-control-exposed participants under the primary ro30 rule. Table F4 (table_f4) summarized pooled phenotype prevalence within deletion- and duplication-defined exposure groups for both ND and control interval families. Table F5 (table_f5) summarized key covariate distributions by primary ND exposure status. Descriptive helper functions included n_pct, median_iqr, prevalence_summary, and summarize_numeric_among_exposed.

**Regression modeling**
Regression modeling was performed in Section G using df_analysis converted to a statsmodels-safe modeling table, df_model_base. The covariate set was explicitly defined in code as COVARIATES = ["age_last_obs",

"log1p_n_unique_visits", "observation_time_days", "pc1", "pc2", "pc3", "pc4", "pc5"], with unavailable columns dropped dynamically if absent from the analytic table. Primary phenotype outcomes were drawn from PHENOTYPE_COLS.

Before model fitting, phenotype and exposure columns were converted to int64 binary variables and covariates to float64. Logistic regression was performed using the helper run_logistic_clean, which:

- selected the outcome, exposure, and covariate columns;
- dropped rows with missing data in any model-required field;
- returned failure status when the modeling subset had zero rows, a constant outcome, or a constant exposure; and
- otherwise fit a statsmodels.Logit model with an intercept and returned sample size, case count, exposed count, odds ratio, 95% confidence interval, and p-value for the exposure term.

The phenotype-level model families were:

- df_g1: primary ND vs none models under nd_any (model = "ND_vs_none_ro30"),
- df_g2: ND vs matched-control models after restricting to participants with either ND or control exposure and excluding overlapping exposure assignments (model = "ND_vs_CTRL_ro30"),
- df_g3: pooled deletion and duplication models using nd_del_any and nd_dup_any,
- df_g4: ND sensitivity-rule models using nd_any_ro50, nd_any_ro70, and nd_any_frac_locus_30.

These outputs were concatenated into df_g_all. Odds-ratio strings were generated in or_ci, and false discovery rate correction was applied across this pooled Section G phenotype-level family using the Benjamini–Hochberg procedure (method = "fdr_bh"), yielding p_fdr and fdr_significant.

The notebook also implemented two higher-order model families. Domain-level pooled models were generated in df_g5_domain, where domain outcomes were derived from dictionary-driven phenotype-to-domain mappings assembled in Section G. These models contrasted pooled ND, pooled deletion, and pooled duplication exposures against unexposed participants, with FDR correction applied within the domain-analysis family only. Multi-domain burden models were generated in df_g6_burden for binary outcomes indicating whether a participant had at least two or at least three affected domains. Readable regression summary tables were then constructed as table_g_summary, table_g_success, table_g_nominal, table_g_fdr, table_g_domain_summary, and table_g_burden_summary.

**Direction-specific analyses**

Section H created mutually informative direction-based groupings from the participant-level exposure table. A direction-specific modeling dataframe, df_dir, was derived from df_model_base, and three binary grouping variables were created:

- DEL_only, defined as nd_del_any == 1 and nd_dup_any == 0,
- DUP_only, defined as nd_dup_any == 1 and nd_del_any == 0,
- NONE, defined as nd_any == 0.

Phenotype prevalence by direction group was summarized in df_h1. Three model families were then fit using run_logistic_clean:

- df_h2: DEL-only versus NONE,
- df_h3: DUP-only versus NONE,
- df_h4: DEL-only versus DUP-only.

**Post-model processing and structure inference**

Following primary model fitting (Sections G–H), all downstream analyses were implemented as structured transformations of the model output tables (df_g_all, df_h_all) and the participant-level analytic dataset (df_model_base / df_analysis). These steps were designed to standardize outputs across model families, enable direct comparison of estimates under alternative analytic conditions, and generate figure-ready datasets corresponding to the primary and supplementary figures.

All model outputs were first restricted to successfully converged fits as determined by the run_logistic_clean helper. Models were excluded if they had zero rows after covariate filtering, constant outcomes, or constant exposure variables. For retained models, odds ratios, 95% confidence intervals, and p-values were formatted into standardized strings (or_ci) and stored alongside raw numeric outputs.
Model outputs from phenotype-level analyses (df_g_all) and direction-specific analyses (df_h_all) were concatenated and annotated with model identifiers, exposure definitions, and phenotype metadata. False discovery rate (FDR) correction was applied within each model family using the Benjamini–Hochberg procedure, and both nominal and FDR-adjusted significance metrics were retained for downstream use.

Figure-ready datasets were derived directly from the harmonized model output tables through filtering, ranking, and annotation steps implemented in pandas. Phenotype-level plots were constructed by selecting outcomes based on nominal significance thresholds and ranking by p-value within model families, with additional inclusion of representative non-significant phenotypes to preserve domain coverage and interpretability. Phenotypes were grouped using dictionary-derived annotations to maintain consistent ordering and labeling across figures. Domain-level and burden-level figures were constructed from aggregated model outputs (df_g5_domain, df_g6_burden), preserving consistent exposure definitions and covariate adjustments. Direction-specific panels were derived from df_h_all, with explicit grouping of DEL-only, DUP-only, and DEL-versus-DUP comparisons.

**Robustness and sensitivity analyses**
Sensitivity analyses addressing potential confounding from somatic structural variation were implemented by excluding structural variant calls flagged as consistent with mosaic chromosomal alterations (mCAs). mCA flags were assigned at the call level within the long-format SV table (ht_sv_calls_long) using heuristic criteria based on variant size, genomic span, and call characteristics. Following mCA flagging, overlap assignment and participant-level exposure construction steps (Sections D–E) were repeated after excluding flagged calls. This resulted in a fully reconstructed exposure table under mCA-filtered conditions, ensuring that downstream analyses reflected re-estimation of carrier definitions rather than post hoc exclusion. The complete regression modeling pipeline (Section G) was then rerun using the filtered dataset, and resulting estimates were aligned with primary results for direct comparison.

To evaluate the contribution of individual loci to observed associations, a leave-one-locus-out (LOLO) framework was implemented. For each curated ND-CNV locus, all overlap assignments corresponding to that locus were excluded from the overlap-collapsed table prior to participant-level aggregation. Exposure variables (nd_any, nd_del_any, nd_dup_any) were then recomputed from the modified overlap table, producing a locus-excluded exposure definition. For each locus-excluded dataset, the full regression modeling pipeline (Section G) was rerun, generating a complete set of phenotype-level association estimates under each exclusion condition. For each phenotype, effect estimates from LOLO models were compared to the corresponding primary model, and summary measures of effect-size shift were computed across loci. These outputs were used to identify loci with the largest influence on model estimates and to construct the LOLO comparison panels.

Robustness analyses were implemented as a coordinated set of re-estimated model families under alternative analytic conditions, including:

- alternative overlap-rule definitions (nd_any_ro50, nd_any_ro70, nd_any_frac_locus_30),
- matched control interval exposures (ctrl_any),
- mCA-filtered exposure definitions, and
- LOLO-derived exposure definitions.

Each condition was implemented by reconstructing participant-level exposure tables and rerunning the full regression modeling pipeline. Resulting model outputs were aggregated and aligned across conditions to enable direct comparison of effect estimates within and across phenotypes. These aligned outputs formed the basis of the multi-panel robustness summaries.

To characterize relationships among neuropsychiatric domains and burden measures, correlation analyses were performed using participant-level domain assignments and derived burden variables. Domain indicators were constructed from phenotype-to-domain mappings defined in Section G, and burden measures included domain count and binary thresholds indicating the presence of at least two or at least three domains. Pairwise Pearson correlation coefficients were computed across all domain and burden variables using the participant-level analytic dataset. Separate correlation matrices were generated for domain-only variables and for combined domain–burden variable sets. These matrices were reshaped into heatmap-ready formats and used to construct the domain and burden correlation visualizations.

**Reproducibility and transfer package**

At completion of all analyses, a structured transfer package was generated containing core analytic datasets and model outputs. Key objects included the participant-level analytic dataset (df_analysis), modeling table (df_model_base), phenotype-level model outputs (df_g_all), direction-specific outputs (df_h_all), and associated phenotype and covariate metadata. Dataframes were exported in both CSV and serialized (pickle) formats using the export_df helper, ensuring compatibility with downstream analyses and figure generation workflows. This export layer preserves full reproducibility of all reported results while separating large-scale data engineering steps from downstream statistical analysis and visualization.

**Global Permutation-Based Enrichment Analysis**

To evaluate whether the aggregate pattern of ND-CNV-associated neuropsychiatric effects exceeded that expected under random exposure assignment, we performed a phenotype-level exposure-permutation analysis using the participant-level modeling dataset generated for the primary phenotype-wide analyses. This analysis was designed to test whether the observed directional consistency across neuropsychiatric phenotypes reflected structured enrichment rather than isolated nominal associations arising through multiple testing. Permutation analyses were conducted separately for pooled ND-CNV carrier status (nd_any), deletion-overlapping ND-CNV carrier status (nd_del_any), and duplication-overlapping ND-CNV carrier status (nd_dup_any). For each exposure definition, participant-level exposure labels were randomly reassigned without replacement while preserving the observed phenotype matrix, phenotype prevalences, sample size, and covariate structure. All phenotype definitions, inclusion criteria, and covariates were identical to those used in the primary phenotype-level logistic regression analyses. Specifically, each permuted dataset was analyzed using the same logistic regression framework incorporating age at last observation (age_last_obs), healthcare utilization (log1p_n_unique_visits), observation time (observation_time_days), and the first five genetic ancestry principal components (PC1–PC5).

For each phenotype-level model, the regression coefficient ($\beta$), standard error (SE), Wald Z-statistic, and P-value associated with the exposure variable were extracted. Aggregate enrichment statistics were then calculated across all modeled neuropsychiatric phenotypes. The primary enrichment statistic was the summed Wald Z-score statistic ($\Sigma Z$), calculated as the aggregate sum of phenotype-level Wald Z-statistics across all modeled neuropsychiatric phenotypes. This statistic preserves both effect direction and effect magnitude while providing a single summary measure of aggregate directional enrichment across the phenotype architecture. Several secondary enrichment metrics were additionally calculated for descriptive and diagnostic purposes. These included: (1) the number and proportion of phenotypes exhibiting positive effect estimates ($\beta > 0$); (2) the summed regression coefficient statistic ($\Sigma\beta$); (3) the mean regression coefficient across phenotypes; (4) the mean Wald Z-statistic; and (5) a signed enrichment score defined as the sum of signed $-\log10(P)$ values, where the sign was determined by the direction of the corresponding regression coefficient. These secondary statistics were evaluated to assess robustness of the observed enrichment pattern across multiple summary frameworks.

A total of 1,000 independent permutations were performed for each exposure definition. For each permutation replicate, the complete set of phenotype-level logistic regression models was re-estimated and aggregate enrichment statistics were recalculated. This procedure generated empirical null distributions representing the expected range of aggregate enrichment statistics under random exposure assignment while preserving the observed phenotype architecture and covariate structure.

Empirical permutation probabilities were calculated by comparing the observed enrichment statistic derived from the original, non-permuted dataset with the corresponding permutation-derived null distribution generated from 1,000 randomized exposure assignments. One-sided empirical probabilities reflected the proportion of permutation replicates producing enrichment statistics equal to or greater than the observed value, whereas two-sided probabilities were calculated relative to the absolute null distribution.

The summed Z-score statistic (ΣZ) was selected as the primary enrichment metric because it directly captures aggregate directional evidence across phenotypes while remaining less sensitive to extreme coefficient estimates arising from sparse outcomes or quasi-complete separation. In contrast, coefficient-based summary statistics may be disproportionately influenced by rare phenotypes with unstable parameter estimates. Accordingly, all primary interpretations of global enrichment were based on ΣZ, whereas secondary enrichment metrics were used as supportive sensitivity analyses.

The purpose of this framework was not to identify additional phenotype-specific associations, but rather to evaluate whether the overall neuropsychiatric architecture associated with ND-CNV carrier status demonstrated greater directional consistency than expected under randomized exposure assignment. Significant deviation from the permutation-derived null distribution was interpreted as evidence of structured enrichment across correlated neuropsychiatric phenotypes.

**Overlap-rule sensitivity analyses and external gnomAD-SV benchmarking**

To evaluate the sensitivity and interval-specificity tradeoffs associated with recurrent neurodevelopmental copy number variant (ND-CNV) assignment, we compared participant carrier yield across alternative overlap definitions within the All of Us Research Program analytic cohort and benchmarked resulting recurrent interval frequencies against external population structural variant resources from gnomAD-SV v2.1.

Observed structural variant (SV) calls were derived from the All of Us Research Program Controlled Tier short-read whole-genome sequencing (srWGS) structural variant resource (CDRv8; GRCh38). The final canonical analytic cohort comprised 71,992 unrelated participants following genomic QC filtering, relatedness pruning, and ancestry integration procedures. Observed SV calls were extracted from chromosome-specific Hail MatrixTables and harmonized into a long-format structural variant table (ht_sv_calls_long) containing participant identifiers, stable observed call identifiers, genomic coordinates, SV type, deletion/duplication indicators, and interval representations. Curated recurrent ND-CNV intervals consisted of 96 deletion- and duplication-specific intervals representing 51 recurrent neurodevelopmental loci harmonized to GRCh38 coordinates. These intervals were assembled from recurrent pathogenic CNV regions represented across prior neurodevelopmental CNV studies, ClinGen dosage-sensitive regions, and established genomic disorder loci. Matched control intervals with similar genomic architectural characteristics were generated separately for specificity analyses but were not included in the external gnomAD benchmarking framework.

Observed SV calls were intersected against curated ND-CNV intervals using explicit overlap metrics computed in Hail, including base-pair overlap, fraction of the observed call overlapped, fraction of the curated interval overlapped, fraction of the smaller interval overlapped, and midpoint inclusion status. These overlap metrics were translated into rule-specific indicators including ro30, ro50, ro70, frac_locus_30, frac_smaller_50, midpoint, and any_overlap. Under the primary analytic framework, participant-level ND-CNV exposure was defined using reciprocal overlap ≥30% (ro30). Sensitivity analyses additionally evaluated stricter reciprocal-overlap thresholds (ro50 and ro70) and a fractional locus-overlap framework (frac_locus_30). Direction matching was enforced throughout all overlap operations such that deletion intervals were evaluated only against deletion calls and duplication intervals only against duplication calls. Carrier-yield summaries were generated across overlap definitions to evaluate the effect of increasing interval stringency on participant assignment and retained locus representation. The ro30 framework was selected as the primary analytic definition because it balanced retention of canonical recurrent ND-CNV loci while excluding weakly overlapping or minimally concordant structural events.

External benchmarking analyses used the Genome Aggregation Database structural variant resource (gnomAD-SV v2.1), a population-scale whole-genome structural variant reference resource containing approximately 14,891 genomes overall. Benchmarking analyses specifically used the publicly distributed BED-compatible interval resource gnomad_v2.1_sv.sites.bed.gz, which contains genomic interval representations and allele-frequency annotations for structural variants identified across the gnomAD-SV call set. Extracted fields included chromosome, genomic coordinates, structural variant identifier, SV type, overall allele frequency (AF), and non-neuro subset allele frequency (NON_NEURO_AF). Benchmarking analyses retained only deletion and duplication events, and primary visualizations preferentially used the NON_NEURO_AF field to reduce potential enrichment from neurologic and psychiatric disease cohorts represented within the broader gnomAD-SV resource.

The gnomAD-SV benchmarking workflow was performed locally using BED-style interval intersection operations implemented in Python and pandas following decompression of the gnomAD-SV BED resource. For each autosome, curated recurrent ND-CNV intervals were compared against chromosome-matched gnomAD-SV intervals filtered to deletion and duplication events. Candidate interval-overlap pairs were identified using genomic interval boundary comparisons, after which reciprocal-overlap metrics and overlap-rule eligibility were computed for each candidate pair. Direction matching was enforced throughout the benchmarking workflow such that deletion intervals were compared only against deletion events and duplication intervals only against duplication events. Unlike the primary All of Us overlap-sensitivity analyses, external benchmarking analyses evaluated only the reciprocal-overlap definitions ro30 and ro50. The stricter ro70, midpoint, and fractional-overlap frameworks were not included in the gnomAD benchmarking analysis. For each recurrent ND-CNV locus, the maximum overlapping gnomAD-SV allele frequency identified under the corresponding reciprocal-overlap threshold was retained for contextual visualization and comparison against All of Us locus prevalence estimates.

The gnomAD-SV comparison framework was intended as contextual population-frequency benchmarking rather than direct replication analysis. Structural variant ascertainment pipelines, sequencing depth, interval representations, quality-control procedures, ancestry composition, and cohort ascertainment differed substantially between All of Us and gnomAD-SV resources. Consequently, differences in observed locus frequencies between datasets should not be interpreted as direct evidence of biological discordance. Increasing reciprocal-overlap stringency preferentially reduced retention of structurally heterogeneous or broad-breakpoint loci, particularly recurrent CNV regions characterized by nested interval architectures. Consistent with increased interval specificity, the ro50 framework generally retained fewer overlapping gnomAD-SV events and produced lower matched allele-frequency estimates than the more permissive ro30 definition.

**Literature concordance and qualitative domain comparison**
To evaluate whether ND-CNV enrichment patterns observed within the All of Us Research Program were directionally consistent with prior neurodevelopmental CNV literature, we performed a qualitative locus-domain concordance analysis across recurrent ND-CNV loci and major psychiatric/neurodevelopmental phenotype domains. Relevant studies were identified from prior large-scale CNV investigations, including clinically ascertained cohorts, neurodevelopmental disorder registries, schizophrenia-focused CNV studies, population-biobank analyses, and cross-disorder CNV resources. Curated references included studies from the Psychiatric Genomics Consortium, UK Biobank analyses, recurrent CNV syndrome cohorts, TOPMed structural variation resources, and large neurodevelopmental CNV aggregation studies. Phenotypic findings were grouped into broad clinically interpretable domains, including cognitive/neurodevelopmental, psychotic, mood/anxiety-related, and broader psychiatric phenotypes.

For each recurrent ND-CNV locus, phenotype-domain relationships reported in the literature were manually reviewed and assigned qualitative evidence scores using the following framework:

- **0** = no clear or consistently reported association
- **1** = reported, variable, or modest association
- **2** = strongly recurrent, canonical, or consistently replicated association

Because prior studies varied substantially in ascertainment strategy, cohort composition, phenotype definition, and statistical methodology, these scores were interpreted qualitatively rather than quantitatively and were not intended to represent effect-size harmonization or formal evidence weighting.

Within All of Us, locus-specific association results were derived from participant-level ND-CNV exposure models using the primary reciprocal-overlap assignment definition (ro30). The AoU-derived scores were intended to summarize broad directional consistency across phenotype domains rather than establish definitive locus-specific replication. For each recurrent locus and phenotype domain combination, directional enrichment patterns were summarized from downstream logistic regression analyses and assigned qualitative directional scores:

- **0** = no directional enrichment observed
- **1** = odds ratio > 1 without nominal statistical significance
- **2** = odds ratio > 1 with nominal $P < 0.05$

**ND-CNV pathway convergence analysis**

To explore potential functional convergence across recurrent neurodevelopmental copy number variant (ND-CNV) loci, we performed an interval-to-gene mapping and pathway enrichment analysis using curated recurrent ND-CNV intervals harmonized to GRCh38 coordinates. This analysis was designed as an exploratory mechanistic-context framework intended to evaluate whether genes captured by recurrent ND-CNV loci demonstrated convergence across biologically interpretable pathways beyond that expected from matched genomic architecture alone.

Curated ND-CNV intervals were derived from the primary recurrent locus resource used throughout the study and included both deletion and duplication intervals. Exact duplicate interval coordinates were collapsed prior to pathway mapping to avoid overrepresentation of recurrent intervals with identical genomic boundaries. Matched genomic control intervals generated for the primary specificity analyses were also included as a comparison framework. These control intervals were selected to approximate recurrent ND-CNV loci with respect to genomic architecture and segmental duplication characteristics while lacking known neurodevelopmental enrichment.

Protein-coding gene annotations were derived from GENCODE v38 (gencode.v38.annotation.gff3). Gene mapping was performed using direct interval overlap between recurrent ND-CNV intervals and annotated gene bodies. Primary analyses used strict interval-gene overlap, while sensitivity analyses additionally evaluated ±100 kb padded interval windows to account for nearby regulatory and breakpoint-adjacent genes. Gene symbols overlapping recurrent ND-CNV intervals and matched control intervals were exported separately for downstream enrichment analyses.

Pathway enrichment analyses were performed using gseapy with the Enrichr framework. Evaluated annotation libraries included: GO Biological Process 2023, Reactome 2022, Human Phenotype Ontology, SynGO 2022, and DisGeNET. Enrichment analyses were conducted independently for ND-CNV interval gene sets and matched-control interval gene sets. For each annotation term, pathway convergence was subsequently evaluated relative to matched genomic controls by comparing the number of ND-CNV-associated genes represented within a pathway against the number of genes contributed by matched control intervals. Odds ratios, Fisher's exact test P values, and false-discovery-rate (FDR) adjusted P values were calculated for pathway enrichment relative to control-derived expectations.

Primary visualizations and manuscript summary tables focused on the top control-adjusted pathways ranked by enrichment significance and effect size. Because recurrent ND-CNV loci span large genomic intervals containing heterogeneous gene content and variable breakpoint architectures, this analysis was intended as an exploratory functional-context framework rather than definitive causal gene prioritization. Consequently, pathway enrichments should be interpreted cautiously and primarily as evidence of broad biological convergence across recurrent neurodevelopmental CNV regions.

### IV. Supplemental Figures

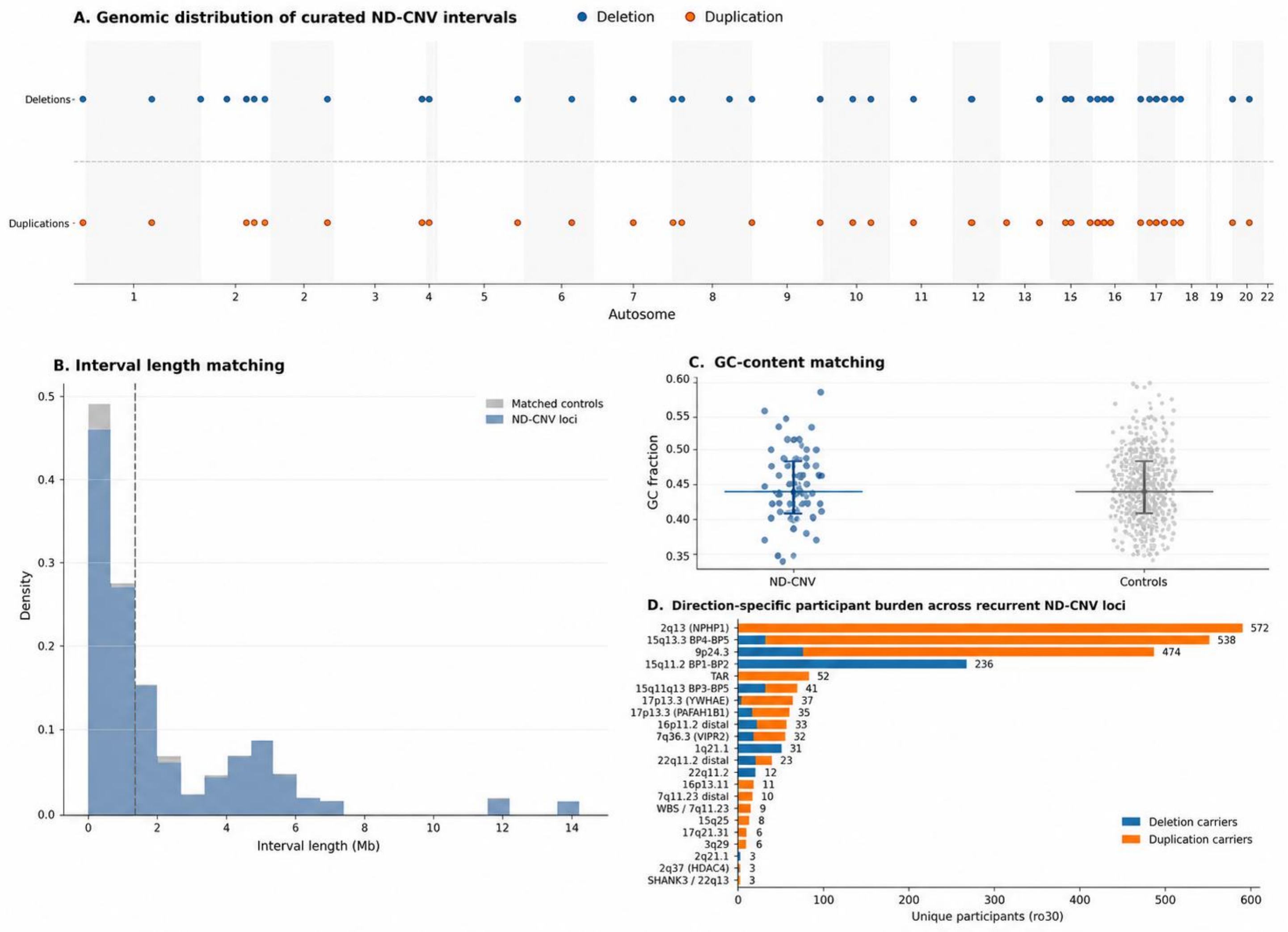


**Figure S1. Genomic architecture and matching characteristics of ND-CNV intervals.**
**(A)** Genomic distribution of curated neurodevelopmental copy number variant (ND-CNV) intervals across autosomes, spanning 96 intervals across 51 loci. Intervals are shown by chromosomal position and stratified by deletion and duplication events. **(B)** Distribution of interval lengths for ND-CNV loci compared with matched control intervals, demonstrating close matching in size. **(C)** Comparison of genomic features between ND-CNV and control intervals, including GC content, confirming similarity across key sequence characteristics. Control intervals were also matched to chromosome and by segmental duplication overlap (not shown). **(D)** Distribution of carrier counts per ND-CNV locus, illustrating the range of locus frequencies and indicating that observed associations are not driven solely by a small number of high-frequency loci.

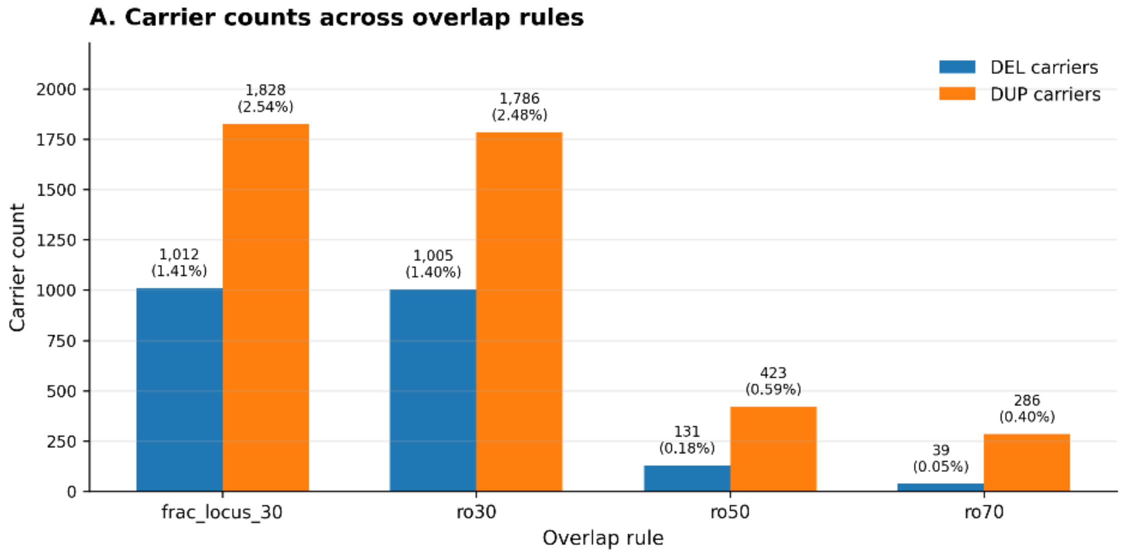
A. Carrier counts across overlap rules
DEL carriers
DUP carriers
Carrier count
Overlap rule
frac_locus_30
ro30
ro50
ro70
1,012 (1.41%)
1,828 (2.54%)
1,005 (1.40%)
1,786 (2.48%)
131 (0.18%)
423 (0.59%)
39 (0.05%)
286 (0.40%)

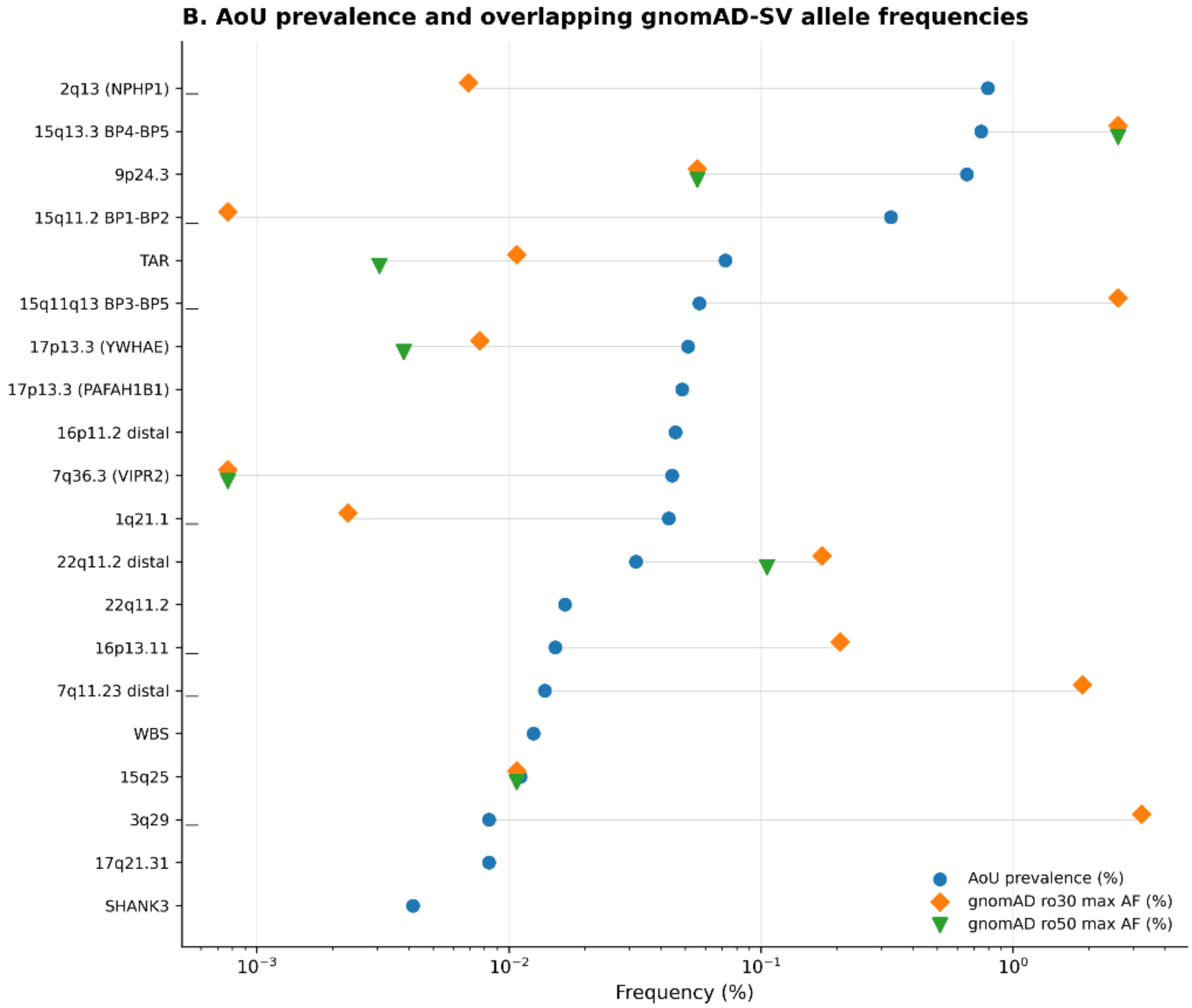
B. AoU prevalence and overlapping gnomAD-SV allele frequencies
2q13 (NPHP1)
15q13.3 BP4-BP5
9p24.3
15q11.2 BP1-BP2
TAR
15q11q13 BP3-BP5
17p13.3 (YWHAE)
17p13.3 (PAFAH1B1)
16p11.2 distal
7q36.3 (VIPR2)
1q21.1
22q11.2 distal
22q11.2
16p13.11
7q11.23 distal
WBS
15q25
3q29
17q21.31
SHANK3
AoU prevalence (%)
gnomAD ro30 max AF (%)
gnomAD ro50 max AF (%)
Frequency (%)

**Figure S2. Overlap-rule sensitivity and external benchmarking of recurrent ND-CNV interval assignment. (A)** Carrier counts across overlap definitions used to assign structural variant calls to curated recurrent neurodevelopmental CNV (ND-CNV) loci in the All of Us cohort. Overlap rules included a permissive locus-fraction criterion (frac_locus_30) and reciprocal-overlap thresholds of 30% (ro30), 50% (ro50), and 70% (ro70). Bars indicate the number of unique deletion (DEL) and duplication (DUP) carriers identified under each rule, with prevalence estimates shown relative to the analytic cohort (N = 71,992). As overlap stringency increased, carrier counts decreased substantially, particularly for deletions, demonstrating the expected tradeoff between sensitivity and interval specificity. The ro30 definition was selected as the primary analytic threshold because it balanced retention of recurrent pathogenic loci with exclusion of minimally overlapping events. **(B)** Comparison of All of Us ND-CNV carrier prevalence estimates with overlapping structural variant frequencies observed in 10,738 whole genomes from unrelated individuals in gnomAD-SV (v2.1, NON_NEURO). Blue circles represent All of Us carrier prevalence estimates for each recurrent locus under the primary ro30 definition. Orange diamonds and green triangles indicate the maximum overlapping gnomAD-SV allele frequencies identified under ro30 and ro50 reciprocal-overlap matching, respectively. Horizontal connector lines link estimates for the same locus across datasets and overlap definitions. Increasing reciprocal-overlap stringency reduced the number of overlapping gnomAD-SV calls and shifted detected frequencies downward, particularly for larger or structurally heterogeneous loci, supporting the use of reciprocal-overlap filtering to improve interval specificity and contextualize observed population prevalence patterns.

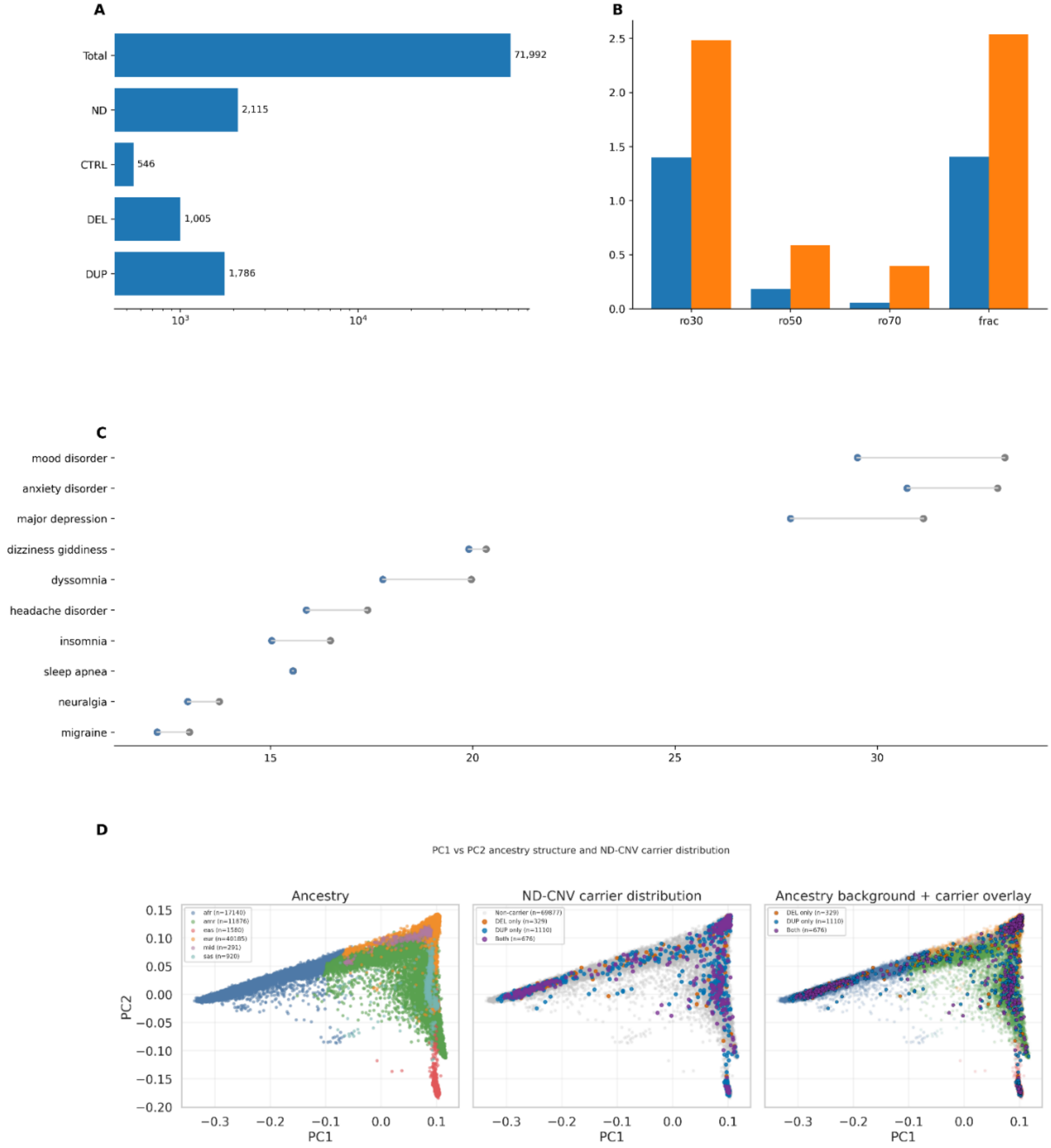
A
Total
ND
CTRL
DEL
DUP
71,992
2,115
546
1,005
1,786
$10^3$
$10^4$
B
ro30
ro50
ro70
frac
C
mood disorder
anxiety disorder
major depression
dizziness giddiness
dyssomnia
headache disorder
insomnia
sleep apnea
neuralgia
migraine
D
PC1 vs PC2 ancestry structure and ND-CNV carrier distribution
Ancestry
afr (n=17140)
amr (n=11876)
eas (n=1580)
eur (n=40185)
mid (n=291)
sas (n=920)
ND-CNV carrier distribution
Non-carrier (n=69877)
DEL only (n=329)
DUP only (n=1110)
Both (n=676)
Ancestry background + carrier overlay
PC1
PC2

**Figure S3. Cohort composition, ND-CNV exposure structure, and genetic ancestry in the analytic dataset. (A)** Composition of the analytic cohort, including total participants, pooled ND-CNV carriers identified under the primary reciprocal-overlap assignment framework (ro30), matched non-carriers, and direction-specific exposure groups consisting of deletion-overlapping and duplication-overlapping ND-CNV carriers. Counts are displayed on a logarithmic scale to accommodate differences in group size. **(B)** Prevalence of deletion-overlapping and duplication-overlapping ND-CNV carriers across alternative overlap-assignment frameworks (ro30, ro50, ro70, and fractional locus coverage). Exposure prevalence is shown separately for deletion-overlapping and duplication-overlapping carriers, illustrating the reduction in carrier prevalence with increasing overlap stringency and the relative stability of fractional locus-based assignment. **(C)** Prevalence of selected clinical phenotypes among pooled ND-CNV carriers and matched non-carriers under the primary ro30 assignment framework. Each phenotype is represented by paired points connected by a line, indicating prevalence among ND-CNV carriers and non-carriers. Phenotypes shown represent the most prevalent conditions in the cohort and are included to provide context regarding baseline clinical burden rather than to support association inference. **(D)** Genetic ancestry structure of the analytic cohort based on principal component analysis (PC1 versus PC2). Individuals are colored according to predicted genetic ancestry, with pooled ND-CNV carriers overlaid to demonstrate their distribution across ancestry clusters. The absence of ancestry-specific clustering among ND-CNV carriers indicates broad representation across ancestry groups and supports inclusion of ancestry principal components as covariates in downstream analyses.

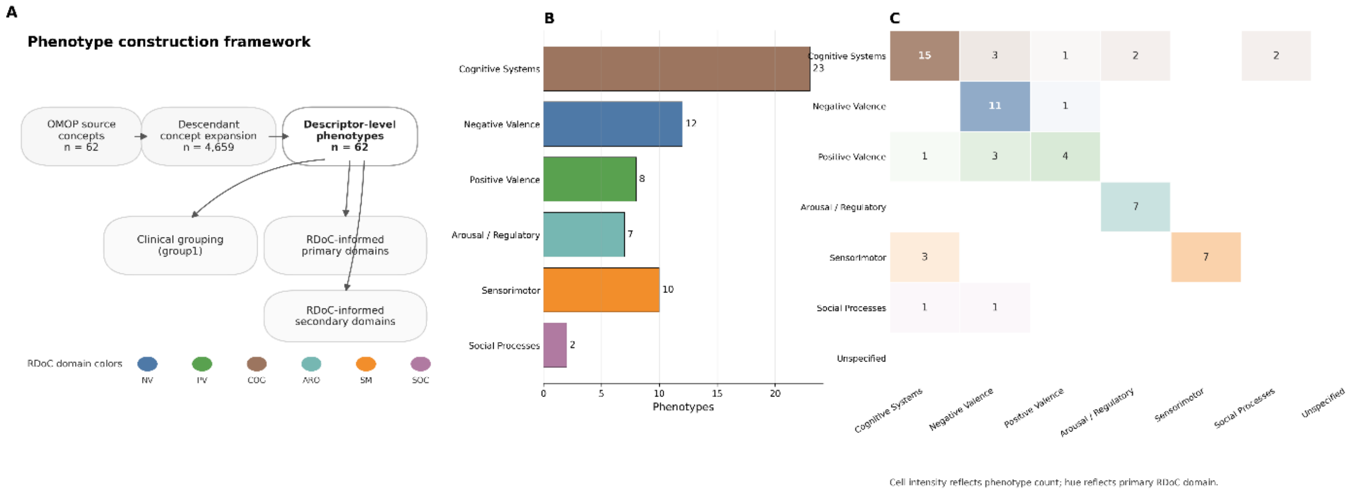


**Figure S4. Construction and domain organization of the phenotype dictionary used for ND-CNV association analyses. (A)** Schematic overview of phenotype construction. Source clinical concepts were derived from OMOP condition concept identifiers and expanded to include all descendant concepts within the OMOP hierarchy. These aggregated concept sets were collapsed into descriptor-level phenotypes representing clinically interpretable disease entities. Each phenotype was subsequently assigned to a clinical grouping (*group1*) and mapped to Research Domain Criteria (RDoC)–informed domains, including both primary and secondary domain assignments. **(B)** Distribution of descriptor-level phenotypes across primary RDoC domains. Each bar represents the number of phenotypes assigned to a given primary domain, with colors corresponding to domain categories (Negative Valence, Positive Valence, Cognitive Systems, Social Processes, Arousal/Regulatory Systems, Sensorimotor Systems, and Unspecified). This distribution reflects the breadth of clinical representation across neuropsychiatric and neurologic domains in the analytic phenotype set. **(C)** Cross-domain mapping between primary and secondary RDoC assignments. Each cell represents the number of phenotypes assigned to a given pair of primary (rows) and secondary (columns) domains. Cell color intensity scales with phenotype count, while hue reflects the primary domain category. This structure illustrates the extent of domain overlap and multi-domain representation inherent to clinically defined phenotypes, supporting downstream analyses that account for pleiotropy and cross-domain effects.

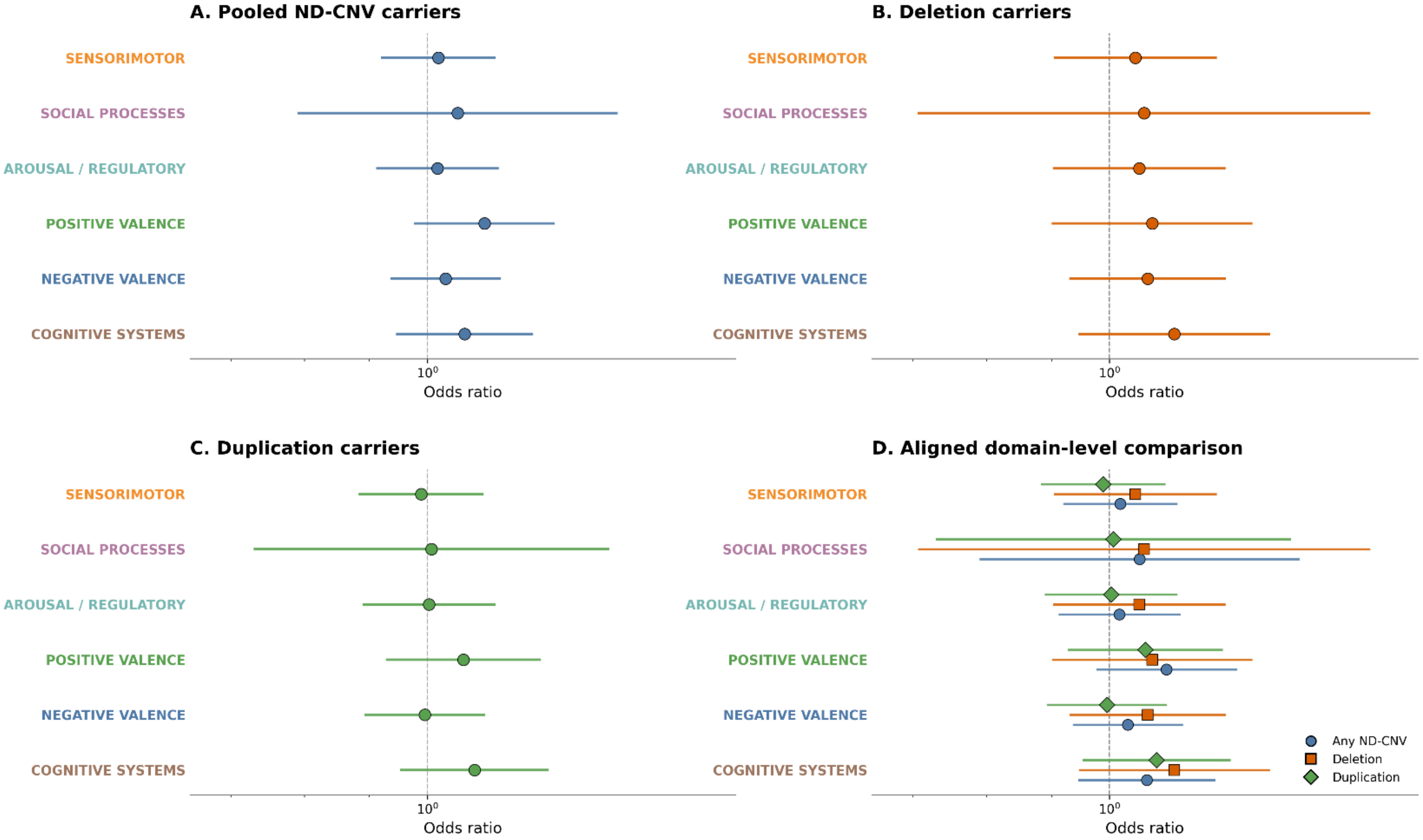


**Figure S5. Domain-level neuropsychiatric associations across ND-CNV exposure models.** Adjusted odds ratios (ORs) and 95% confidence intervals (CIs) for associations between neurodevelopmental copy number variant (ND-CNV) exposure status and neuropsychiatric domains defined using the Research Domain Criteria (RDoC) framework. Domain assignments were derived from phenotype-level mappings and reflect non-mutually exclusive classification across six functional systems: Cognitive Systems (COG), Negative Valence Systems (NV), Positive Valence Systems (PV), Arousal/Regulatory Systems (ARO), Social Processes (SOC), and Sensorimotor Systems (SM). **(A)** Pooled ND-CNV carriers versus non-carriers. Across domains, effect estimates are modest and generally oriented above the null (OR = 1), with the largest nominal signal observed for Positive Valence Systems (PV). **(B)** Deletion-overlapping ND-CNV carriers versus non-carriers. Deletion-overlapping carriers demonstrate somewhat stronger and more broadly distributed enrichment across domains, particularly within Cognitive Systems (COG) and Positive Valence Systems (PV). **(C)** Duplication-overlapping ND-CNV carriers versus non-carriers. Duplication-overlapping carrier effects are generally smaller and closer to the null, with no domain showing substantial deviation. **(D)** Aligned comparison across exposure models. Domain-level estimates for pooled ND-CNV, deletion-overlapping ND-CNV, and duplication-overlapping ND-CNV exposure models are displayed together to facilitate direct comparison. Across domains, effect estimates are generally concordant in direction but differ modestly in magnitude, with deletion-overlapping models tending toward larger effect estimates than duplication-overlapping models. All models were estimated using logistic regression adjusted for age at last observation, log-transformed healthcare utilization (unique clinical visits), observation time, and genetic ancestry principal components (PC1–PC5). Points represent ORs and horizontal lines represent 95% confidence intervals; the dashed vertical line indicates OR = 1. False discovery rate (FDR) correction was applied within the domain-level model family, and no associations met the FDR-significance threshold.

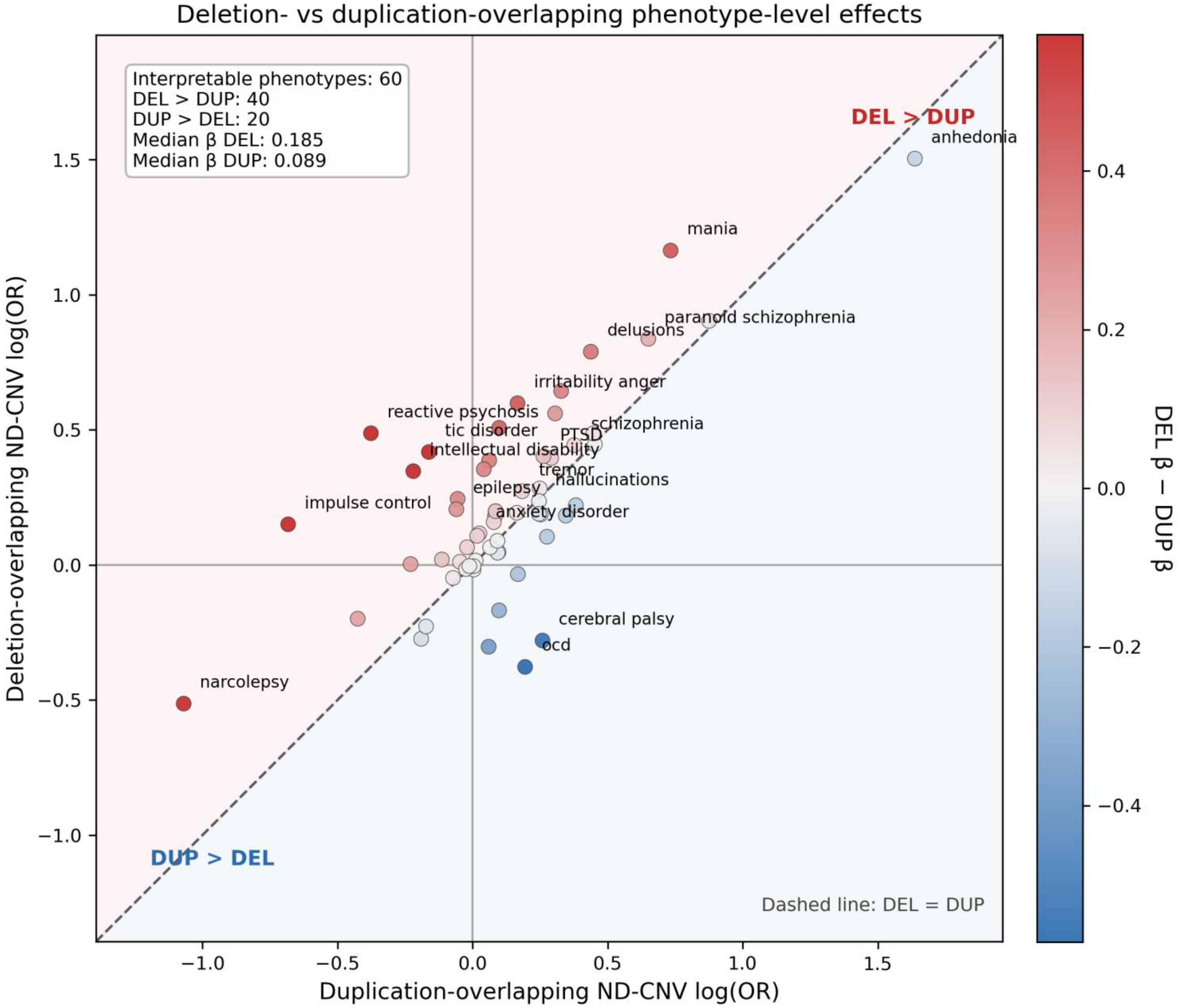


**Figure S6. Deletion-overlapping and duplication-overlapping ND-CNV effect architecture across neuropsychiatric phenotypes.** Comparison of deletion-overlapping and duplication-overlapping ND-CNV effect estimates across phenotype-level association models. Each point represents one neuropsychiatric phenotype. The diagonal line indicates equivalent effect estimates between deletion-overlapping and duplication-overlapping carrier models. Points above the diagonal indicate stronger deletion-overlapping effects, whereas points below the diagonal indicate stronger duplication-overlapping effects. Analyses were restricted to interpretable regression estimates to avoid complete-separation artifacts. Overall, deletion-overlapping carrier models demonstrated larger effect estimates across a greater proportion of phenotypes, supporting direction-specific heterogeneity in ND-CNV-associated neuropsychiatric liability.

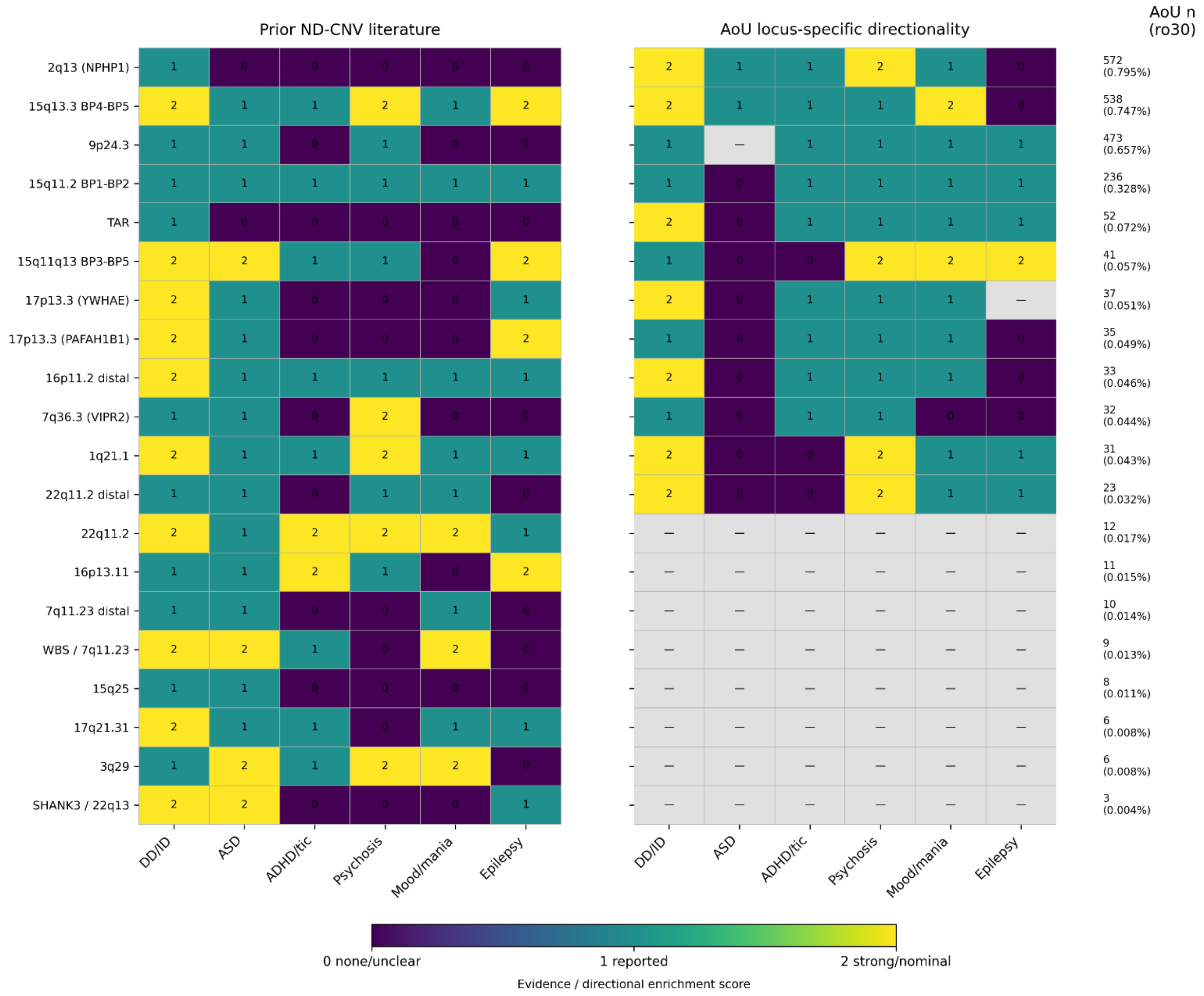


**Figure S7. Qualitative concordance of recurrent ND-CNV phenotypic domains in prior literature and All of Us.** Heatmaps compare manually curated literature-supported phenotypic domains for recurrent ND-CNV loci with locus-specific directional enrichment patterns observed in the All of Us Research Program. Literature scores indicate whether a phenotype-domain association was not clearly established/unclear (0), reported or variably supported (1), or strongly recurrent/canonical (2) based on prior CNV studies. AoU scores indicate no directional enrichment (0), directional enrichment with OR > 1 (1), or OR > 1 with nominal P < 0.05 (2). Rightmost values show ro30 carrier counts and prevalence estimates within the AoU analytic cohort. Grey cells in the AoU panel indicate loci or phenotype-domain combinations that were not evaluable or were suppressed under All of Us privacy-policy enforcement thresholds for low cell counts and protected output rules; these cells should not be interpreted as evidence of absent association or negative findings. Literature directional/domain assignments were manually curated from prior large-scale CNV studies, registry cohorts, population biobanks, and cross-disorder neurodevelopmental CNV analyses, including Birnbaum et al. (2022), Curran et al. (2013), Moreno-De-Luca et al. (2013), Hoeffding et al. (2017), Kendall et al. (2019), Marshall et al. (2017), Mitrakos et al. (2024), Pollak et al. (2019), Thygesen et al. (2021), Zarrei et al. (2019), and TOPMed structural variation reference analyses (Jun et al., 2023).

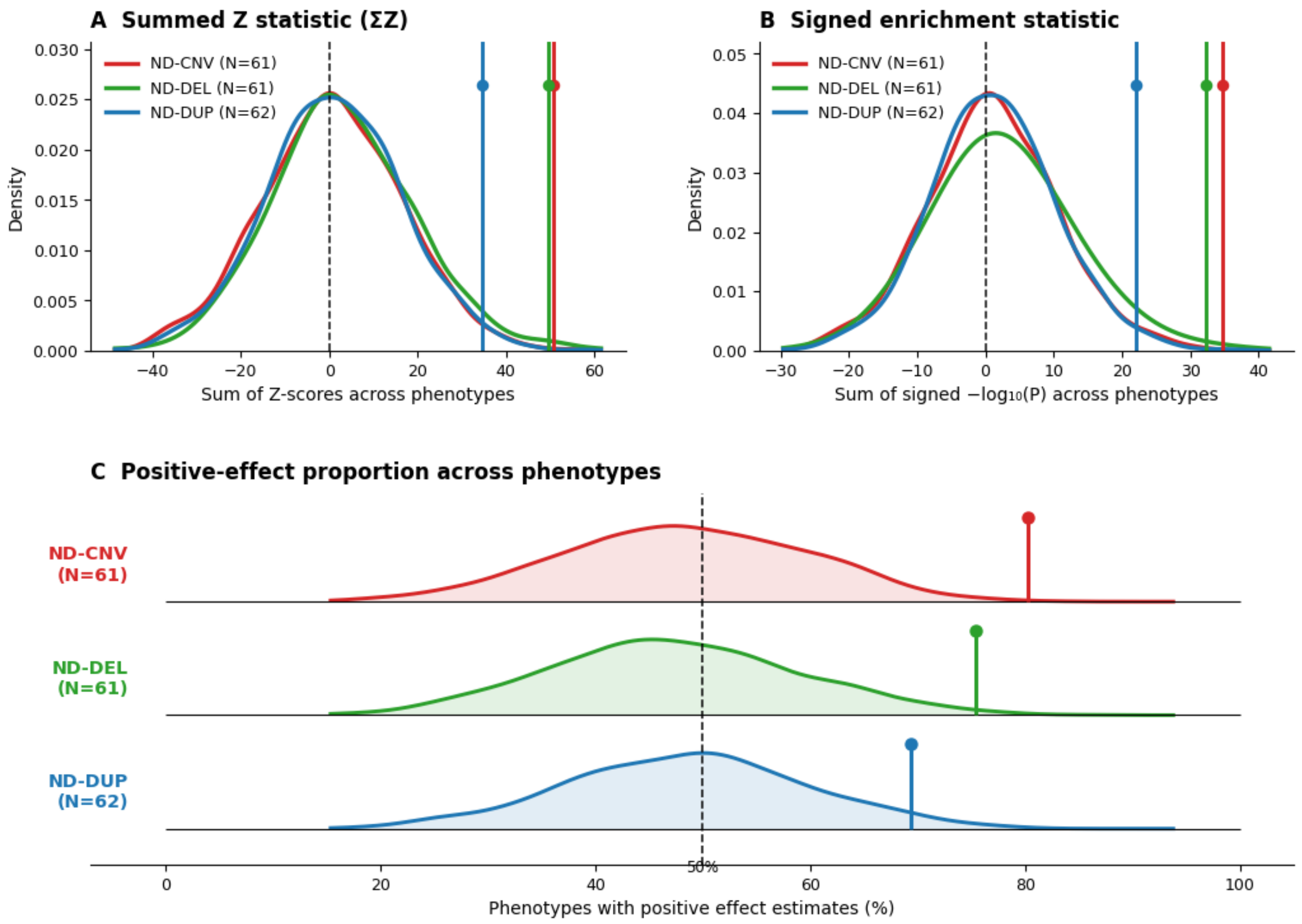


**Figure S8. Sensitivity of global permutation enrichment results across alternative summary metrics.** Permutation-based global enrichment analyses were repeated using complementary aggregate statistics to evaluate whether the observed ND-CNV-associated neuropsychiatric signal was robust to the choice of enrichment metric. **(A)** Empirical null distributions for the summed Wald Z-score statistic (ΣZ) across phenotype-level logistic regression models. Observed values were ΣZ = 50.85 for pooled ND-CNV carriers, ΣZ = 49.67 for deletion-overlapping ND-CNV carriers, and ΣZ = 34.62 for duplication-overlapping ND-CNV carriers, corresponding to empirical permutation probabilities of P = 0.002, P = 0.008, and P = 0.016, respectively. **(B)** Empirical null distributions for the signed enrichment statistic, calculated as the sum of signed −log10(P) values across phenotype-level models. Observed signed enrichment values were 34.66 for pooled ND-CNV carriers, 32.23 for deletion-overlapping ND-CNV carriers, and 21.98 for duplication-overlapping ND-CNV carriers, corresponding to empirical permutation probabilities of P = 0.002, P = 0.013, and P = 0.018, respectively. **(C)** Empirical null distributions for the proportion of phenotype-level associations with positive effect estimates. Observed positive-effect proportions were 80.3% for pooled ND-CNV carriers (49/61 phenotypes), 75.4% for deletion-overlapping ND-CNV carriers (46/61 phenotypes), and 69.4% for duplication-overlapping ND-CNV carriers (43/62 phenotypes), corresponding to empirical permutation probabilities of P = 0.002, P = 0.011, and P = 0.039, respectively. Across all three summary metrics, observed enrichment exceeded expectations under randomized exposure assignment, supporting the robustness of the global enrichment signal.

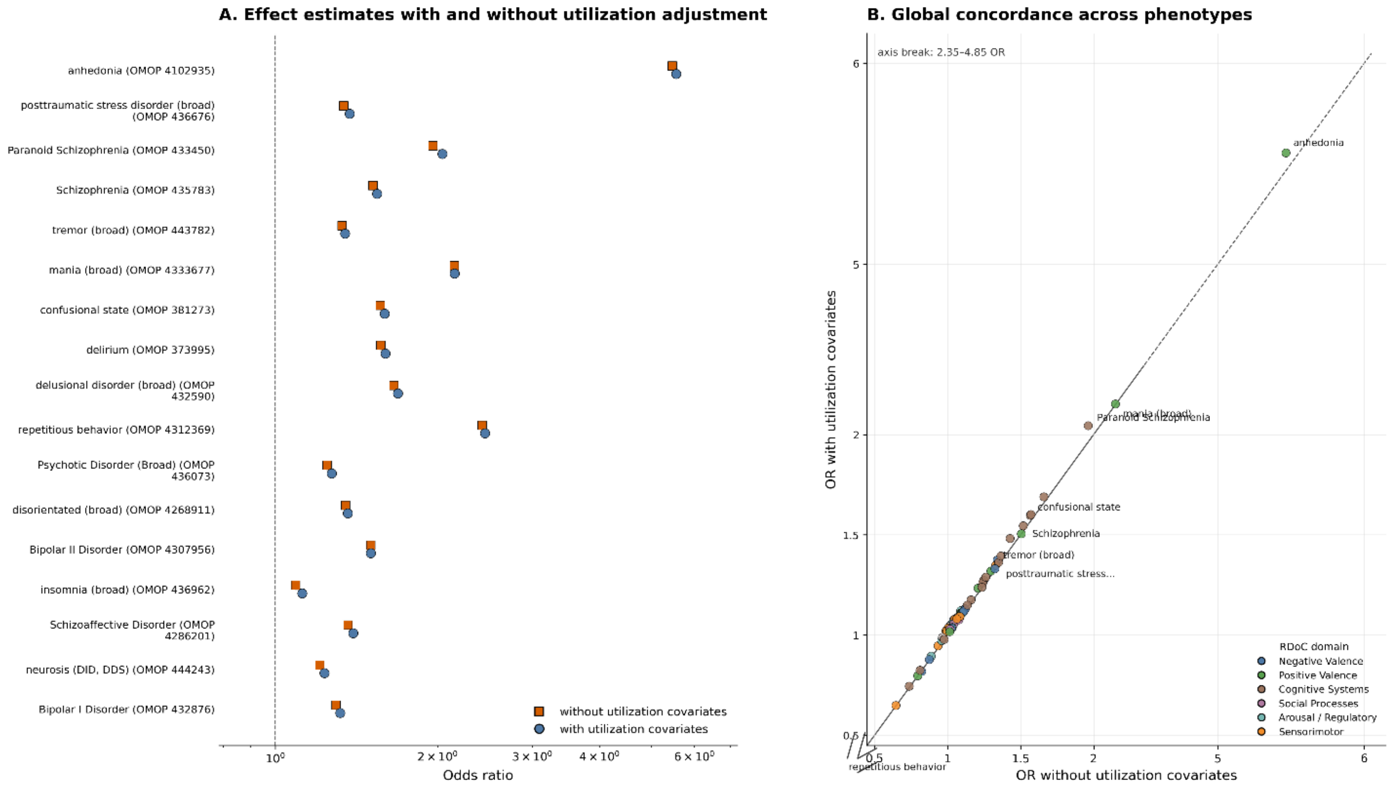


**Figure S9. Sensitivity of ND-CNV associations to healthcare utilization and ascertainment. (A)** Comparison of effect size estimates for selected phenotypes with and without adjustment for healthcare utilization covariates. Odds ratios (ORs) are shown for models excluding (orange squares) and including (blue circles) measures of healthcare utilization, including visit density and observation time. Lines connect paired estimates for each phenotype. Phenotypes include all nominally significant associations under the fully adjusted model ($P < 0.05$), along with a subset of additional phenotypes selected based on significance ranking. Effect estimates are shown on a logarithmic scale, with the vertical dashed line indicating OR = 1. **(B)** Concordance of effect size estimates across all tested phenotypes before and after adjustment for healthcare utilization. Each point represents a phenotype, colored by primary RDoC domain. The dashed diagonal line indicates equality between models. To improve visualization of most phenotypes clustered at modest effect sizes, a scale break is applied to both axes (OR 2.35–4.85), compressing the upper range while preserving relative ordering. Selected phenotypes with the strongest associations are labeled. The tight clustering of points along the identity line indicates that adjustment for healthcare utilization has minimal impact on estimated effect sizes.

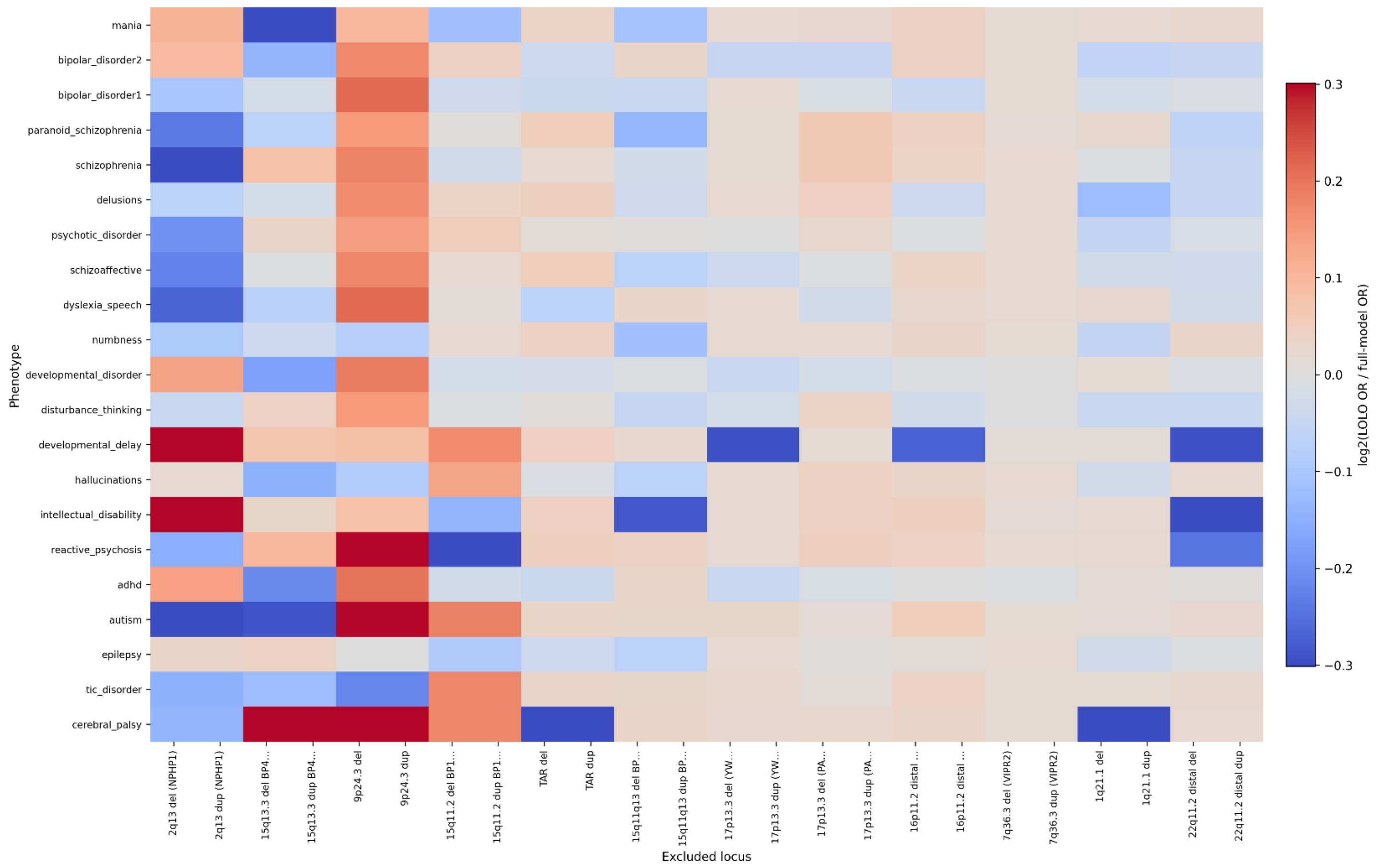


**Figure S10. Stability of ND-CNV associations under leave-one-locus-out (LOLO) analysis.** Heatmap showing the change in effect size for each phenotype following sequential exclusion of individual ND-CNV loci. Each column corresponds to a single locus removed from the carrier definition, and each row corresponds to a phenotype. Cell values represent the $\log_2$-transformed ratio of the LOLO-derived odds ratio (OR) to the full-model OR ($\log_2$[OR_LOLO / OR_full]). Color intensity reflects the magnitude of deviation from the full model, with red indicating an increase in effect size upon locus removal and blue indicating a decrease. Values near zero (white) indicate minimal change. Columns are ordered by the number of excluded carriers per locus. Across phenotypes, effect estimates remain largely stable under LOLO perturbation, with no consistent locus-specific patterns of deviation. The absence of strong vertical banding indicates that observed associations are not driven by any single CNV locus but instead reflect distributed effects across loci.

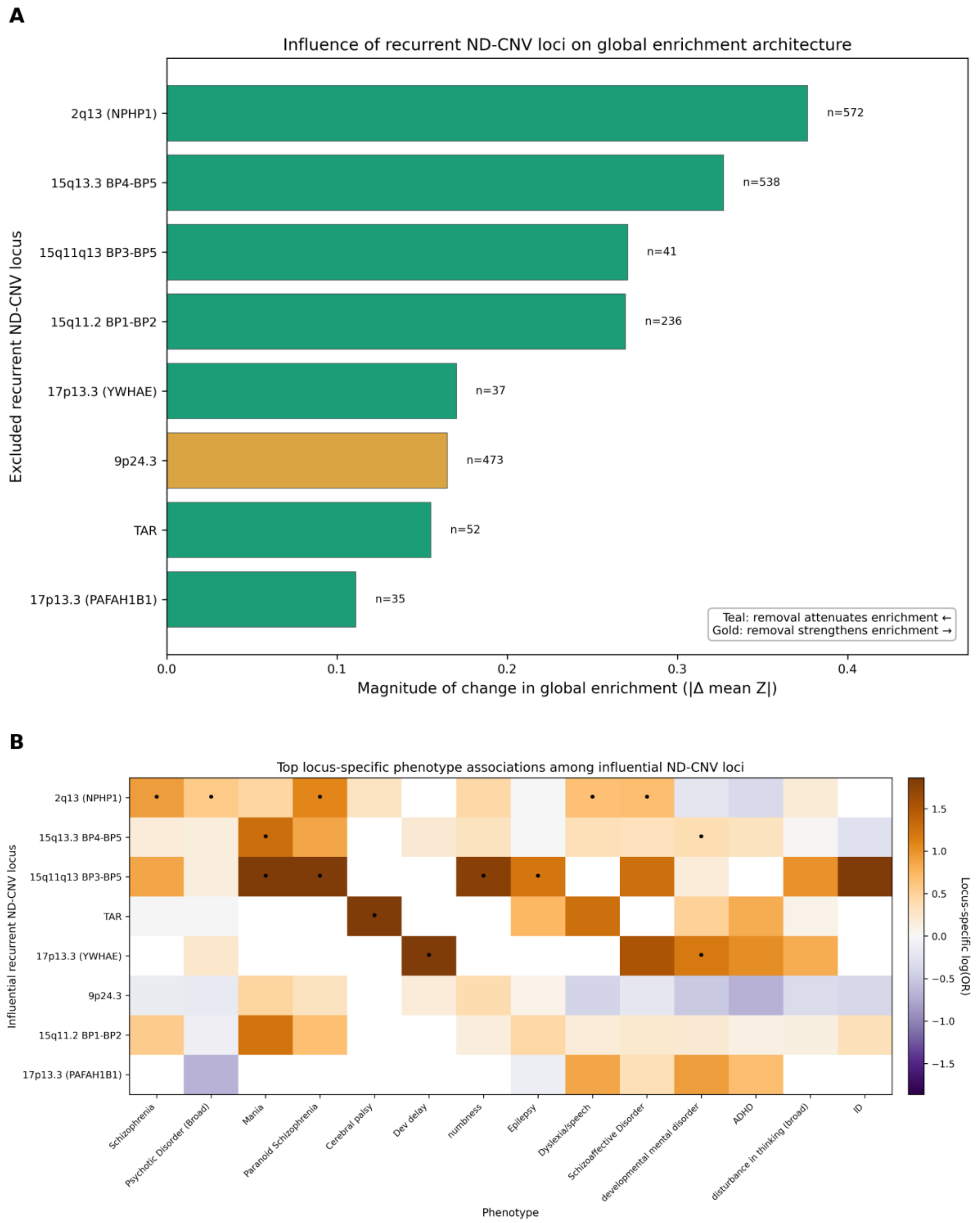


**Figure S11. Locus-level contributions to global ND-CNV neuropsychiatric enrichment.**
(A) Leave-one-locus-out analysis ranking recurrent ND-CNV loci by the magnitude of change in global enrichment after locus exclusion, quantified as |Δ mean Z| across modeled phenotypes. Teal bars indicate loci whose removal attenuated global enrichment, whereas gold bars indicate loci whose removal strengthened global enrichment. (B) Heatmap of top locus-specific phenotype associations among loci prioritized by leave-one-locus-out influence. Cell color represents locus-specific log odds ratios, and dots indicate nominal P < 0.05. Together, these analyses show that pooled ND-CNV-associated neuropsychiatric enrichment reflects contributions from multiple recurrent loci with heterogeneous phenotype-specific association profiles.

### V. Supplemental Tables and Full Pipeline Outputs

To facilitate reproducibility and navigation of the analytical workflow, Supplementary Tables 1 and 2 provide provenance mapping between manuscript figures, exported analytical outputs, and major workspace resources. Supplementary Table 3 contains the external interval, phenotype, and annotation resources used throughout the study.

**Supplemental Table 1. Figure and table provenance map.** Comprehensive provenance mapping for all main and supplemental figures. For each figure, the table identifies the primary source datasets, exported analysis outputs, and source-data resources used in figure generation. This table provides a direct linkage between All of Us workspace outputs, external validation resources, manually curated source-data files where applicable, and the final manuscript figures, facilitating transparency and reproducibility of the analytical workflow.

**Supplemental Table 2. Workspace output inventory and analytical resource catalog.** Inventory of major datasets, intermediate outputs, and exported analytical resources generated throughout the study workflow. Files are organized by pipeline stage, including interval curation, phenotype construction, exposure assignment, association modeling, robustness analyses, correlation analyses, permutation testing, pathway enrichment analyses, and external validation resources. The table serves as a reference guide linking workspace outputs to their role in downstream analyses and manuscript generation.

**Supplemental Table 3. External resources and curated reference datasets.**
This table provides the external and curated resources used in the construction of analytic variables and exposure definitions. Included are (i) the phenotype dictionary with OMOP concept mappings and RDoC-informed domain assignments, (ii) matched negative control genomic intervals (raw and ±100 kb padded) used for specificity analyses, (iii) the ND-CNV interval curation table summarizing genomic coordinates, source annotations, and harmonization procedures, and (iv) the raw and padded BED file externals for ND-CNV interval calling. These resources form the foundational inputs for all downstream analyses.

**Full pipeline outputs for this project have been deposited into Open Science Framework**:

https://osf.io/5dxtv/overview?view_only=4a86da9f04c6456d871ca9cf96e9027b

### VI. All of Us (AoU) and AI Disclosures

**Data governance and privacy compliance**

All analyses were conducted using controlled-tier data from the All of Us Research Program within the Researcher Workbench in accordance with program data use policies and researcher agreements. Access to individual-level data required completion of All of Us Responsible Conduct of Research training and adherence to the Data User Code of Conduct. Analyses were restricted to participants included in the Controlled Data Release version 8 (CDRv8), all of whom provided appropriate consent for the use of their electronic health record and genomic data for research purposes. Data linkage across genomic, phenotypic, and demographic sources was performed within the secure environment using program-approved identifiers.

**Privacy-policy enforcement and low-count suppression**

To comply with All of Us Research Program Controlled Tier privacy and disclosure-protection policies, locus- and phenotype-specific analyses involving small participant counts were subject to output suppression and restricted reporting procedures. In particular, certain locus-domain combinations with low carrier counts, unstable effect estimation, or policy-restricted cell sizes were excluded from exported summary tables and downstream visualization layers. These suppressed combinations are represented as grey cells in selected supplementary figures.

Importantly, suppressed or unavailable cells should not be interpreted as evidence of absent association, null effect, or lack of directional enrichment. Rather, these entries reflect privacy-preserving reporting constraints imposed by the All of Us Researcher Workbench environment. Consequently, qualitative comparisons between loci and phenotype domains should be interpreted within the context of these reporting limitations.

**Reproducibility and data handling**

Analytical workflows were implemented within the All of Us Researcher Workbench using version-controlled notebooks and structured data processing pipelines. Intermediate datasets were stored within the secure workspace and represented using Hail tables, MatrixTables, and tabular formats optimized for large-scale genomic analysis. Exported data products were limited to derived, de-identified summary-level outputs necessary to reproduce reported results.

**Use of computational assistance tools**

Computational tools, including large language models, were used in a limited capacity to assist with code debugging, pipeline refinement, and manuscript preparation. These tools were used to support syntax correction, error interpretation, and refinement of written descriptions of methods and results. No individual-level participant data or protected information were entered into external tools at any stage.

All analytical decisions, study design elements, data processing steps, and statistical analyses were performed and validated by the authors. The use of computational assistance tools did not influence the scientific conclusions of the study.
